# Current status of anti-obesity medications and performance, an EHR based survey

**DOI:** 10.1101/2024.12.02.24318314

**Authors:** Xiaoyang Ruan, Rui Li, Liwei Wang, Shuyu Lu, Andrew Wen, Murali Sameer, Hongfang Liu

**Affiliations:** Department of health data science and AI, McWilliams School of Biomedical Informatics, University of Texas Health Science Center at Houston; Department of Clinical and Health Informatics, McWilliams School of Biomedical Informatics, University of Texas Health Science Center at Houston; Department of Surgery, McGovern Medical School, University of Texas Health Science Center at Houston

**Author notes:** Corresponding author: Hongfang Liu, PhD Postal address: 7000 Fannin Street #Suite 600, Houston, TX 77030.

**Keywords:** Anti-Obesity Medication, Body Weight, BMI, Lipid Profile, EHR

## Abstract

**Background:** Over the past two decades, the Food and Drug Administration (FDA) has significantly increased the approval of anti-obesity medications (AOMs) for obesity management. Both FDA-approved AOMs (F-AOMs) and Off-label AOMs (O-AOMs) have gained popularity and demonstrated promising results in randomized clinical trials (RCTs). However, their effectiveness in real-world settings remains less understood. In this study, we evaluated population-level responses to AOMs and individual variability, leveraging electronic health records (EHRs) as real-world data sources for a comprehensive analysis of obesity relevant metabolic metrics.

**Methods:** EHRs of patients with obesity or overweight diagnosis were retrieved from the University of Texas Physician (UT-Physician) and EPIC COSMOS database. F-AOMs and O-AOMs were analyzed for their effects on obesity relevant metrics including body weight, body mass index (BMI), blood pressure (BP), high-density lipoprotein (HDL), low-density lipoprotein (LDL), HbA1c, and triglyceride levels.

**Results:** From the UT-Physician (U) and COSMOS (C) datasets, we identified 610K (2015-2025) and 3.6M (2018-2024) patients as obese or overweight, and 71,318 (11.7%) and 1M (27.9%) with AOM exposures, respectively. During the study period, [U:72%; C:67%] of patients experienced more than one treatment session. The median exposure durations were [U:2.7; C:2.7] months for FDA-approved AOMs (F-AOMs) and [U:.3.0; C:3.1] months for off-label AOMs (O-AOMs). Across both cohorts, F-AOMs generally demonstrated greater weight-loss effects than O-AOMs. Tirzepatide and semaglutide were the most effective F-AOMs. Tirzepatide achieved 10% weight loss target in [U:42%; C:48%] of patients with long-term exposure (65-66 weeks), while semaglutide achieved this threshold in [U:23%; C:33%] of patients with long-term exposure (70-72 weeks). Both medications were also associated with improvements in blood pressure, HDL, LDL, triglycerides, and HbA1c. Among O-AOMs, topiramate demonstrated the most favorable long-term (84-86 weeks) outcomes, with 13% of patients achieving 10% weight loss in both cohorts. Substantial interindividual variability in treatment response was observed across all AOMs, regardless of diagnosis type or exposure duration. Notably, weight gain was observed for all AOMs, ranging from [U:8%; C:12%] for long-term tirzepatide exposure to [U:57%; C:53%] for long-term lisdexamfetamine exposure. Weight regain following AOM discontinuation was consistently observed across both cohorts for both O-AOMs (e.g., lisdexamfetamine) and F-AOMs (e.g., phentermine w/wo topiramate). Interestingly, the combination of metformin with semaglutide or tirzepatide was associated with attenuated weight regain after treatment discontinuation compared with semaglutide or tirzepatide monotherapy.

**Conclusions:** Our findings demonstrate real-world effectiveness of AOMs, particularly FDA-approved AOMs, in a subset of patients, while revealing substantial interindividual variability and notable discrepancies between EHR-based evidence and clinical trial results. These findings highlight the challenges of translating trial outcomes to routine practice and underscore the need for personalized obesity pharmacotherapy to improve effectiveness, adherence, and long-term sustainability of care.

## Introduction

Obesity is a multifactorial disease that affects over a third of the world’s population ^1,2^ , and ∼34% of adults and 15-20% of children/adolescents in the USA ^3^. Over 85% of adults are projected to be overweight or obese by 2030 in the USA ^4^. Obesity greatly increases the risk of a wide spectrum of chronic diseases ^5^, and causes significant social economic burden ^6,7^. Obesity arises from a complex interplay of genetic ^8,9,10,11^ , epigenetic ^12,13,14,15^, non-genetic factors ^16,17^ , and genetic-environment interactions ^18,19^. They together cause energy imbalance, excessive fat deposition, and obesity phenotype.^12^

During the past decades, a wide variety of therapies have emerged for obesity treatment, such as bariatric surgeries (BSs), anti-obesity medicines (AOMs), hydrogels, vagal nerve blockade, along with traditional dietary control and exercise. However, no single or combination of therapies perform consistently across individual patients, either in short or long term. For example, meta-analysis indicates BS patients have wide interindividual response during longer-term follow-up ^20,21^. A prospective study of 6000 Roux-en-Y gastric bypass patients showed 17.1% with inadequate weight loss 1 year after surgery, and 23.1% as non-responders 5 years after surgery ^22^. In contrast, the majority of AOMs reduce average body weight by less than 10% at tolerable doses, with remarkably wide interindividual variability ^23^. To illustrate, in Orlistat phase 3 trial ^24^, 27% and 59% participants failed to achieve 5% and 10% weight loss target in 1 year. In Phentermine phase 3 trial ^25^, 30% and 52% participants in the high dose (15mg) intervention group failed to achieve 5% and 10% weight loss targets in 1 year. While GLP-1 drug Semaglutide claims to achieve an average of >10% weight loss ^26^, this level of performance was seen in only 69% of 1306 participants ^27^. Even with additional intensive behavioral therapy, 10% weight loss was achieved in less than 75% participants^27^. The latestly approved GLP-1/GIP drug Tirzepatide ^28^ achieved a 10% weight loss target in only 69% participants at a relatively safe dose level (5mg).

Notably, most of the weight loss performance results come from randomized clinical trials (RCTs), which have rigorous exclusion criteria (e.g. comorbidities), study regulations, limited study duration ^29^, and potential conflicts of interest stemming from pharmaceutical industry sponsorship. Because of this, the outcomes are not consistently replicated in real-world settings. Whereas a recent study based on Electronic Health Record (EHR) ^30^ also implemented strict inclusion criteria (e.g. patients in weight management programs with nutritional, behavioral, and psychological guidelines, and two face-to-face visits in at least 3 months of follow-up), which resulted in a very limited cohort size. In view of the shortcomings, here we conducted a comprehensive walk through of the UT-Physician EHR systems on patients’ exposures and responses to various AOMs, and validated some of the findings in a nationwide large cohort curated from the EPIC COSMOS database. We aim to provide a landscape view of the prevalence and effectiveness of AOMs in the general population, along with the complexity of treatment journey experienced by obesity patients, and highlight the desiderata of next generation phenotyping for precision medicine.

## Material and Methods

### EHR databases and study cohorts

#### UT-Physician cohort

The UT-Physician cohort includes 626,078 patients with either diagnosis of obesity (n=365,091) or overweight with comorbidities (n=260,987) from 2015/01/01 to 2025/06/30 in the UT Health EHR system. The system was served by Allscripts before 2020, and then switched to EPIC. From May 2020, the UT EPIC system started to record drug dispensing information reported by more than 50,000 pharmacies across all 50 states and the District of Columbia. The Drug dispense records were used as a proxy for drug exposure. All records were mapped to the Observational Medical Outcomes Partnership (OMOP) common data model ^31^ before further analysis. For obesity we considered six diagnosis subtypes (Supp Text 1). For overweight we considered patients with any comorbidities (Supp Text 2) within 1 year prior to a BMI measurement ≥ 27. After excluding those with obesity-related surgery and without demographic information, finally a total of 610,372 (97.5%) patients were left, among which 71,318 (11.7%) have at least one AOM pharmacy dispense records. The sample processing flow is shown in Figure 1a.

**Figure 1.**
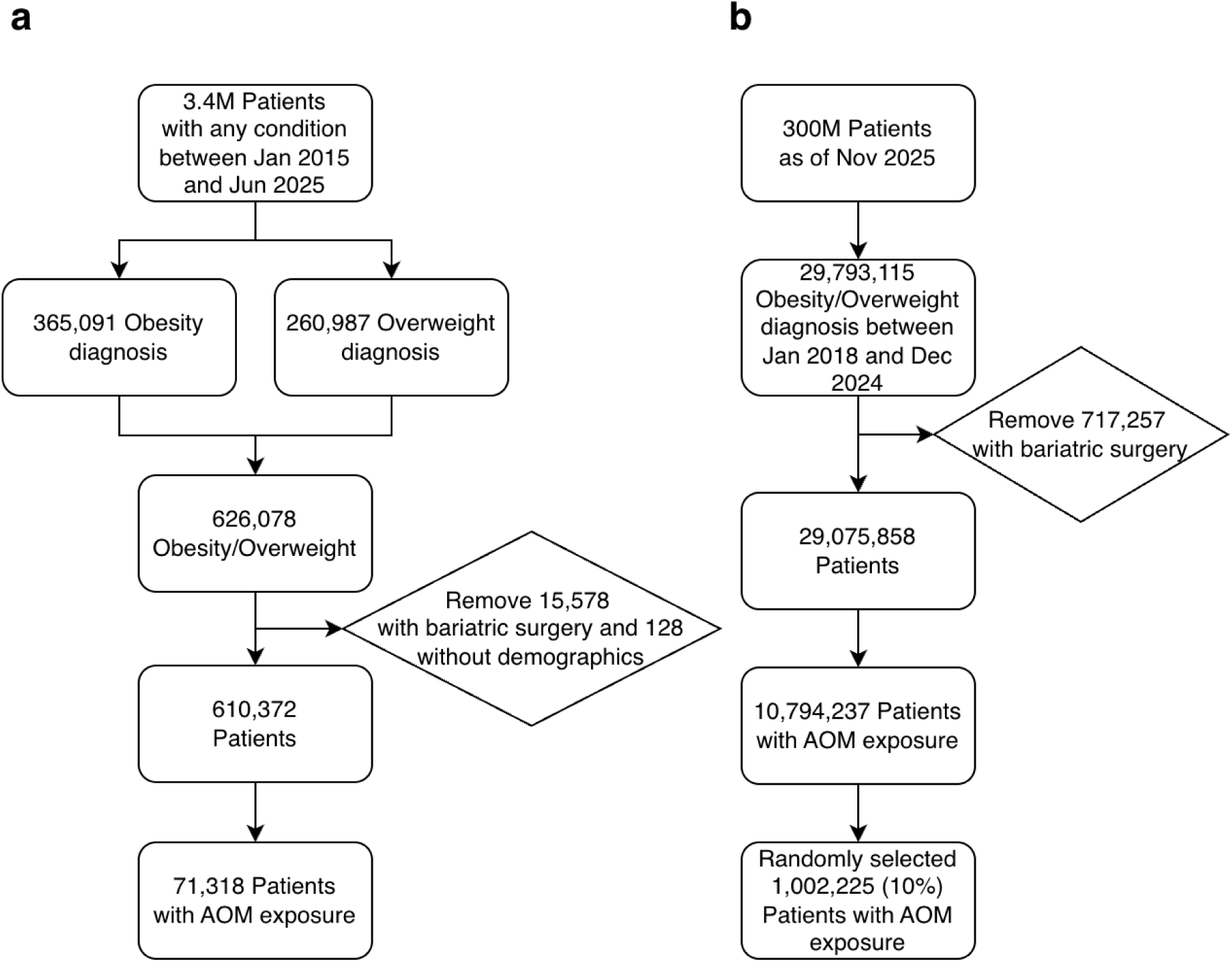
Sample processing flow for a) UT-Physician cohort, b) COSMOS cohort.

#### COSMOS cohort

COSMOS is a platform enabling users to leverage de-identified data from over 300 million patients (as of Nov 2025) from more than 1,800 hospitals across the United States and Lebanon to advance human health through research and clinical integration. COSMOS EHR is based on the caboodle data model of EPIC data warehouse, and has drug dispensing information recorded since Apr 2007. For the COSMOS cohort, we selected 29,793,115 patients with ICD-9 and ICD-10 based obesity diagnosis (Supp Text 3) from 2018/01/01 to 2024/12/31. We further excluded those with any obesity-related surgeries (Supp Text 4), and 29,075,858 (97.6%) patients left. Among which 10,794,237 (37%) were exposed to at least one AOM. Due to limited computation capacity, we randomly selected 1,002,225 (∼10%) patients for downstream analysis. The sample processing flow is shown in Figure 1b.

#### FDA approved AOMs (F-AOMs) and Off-Label AOMs (O-AOMs)

For F-AOMs, we considered bupropion naltrexone, orlistat, phentermine, phentermine topiramate, liraglutide, semaglutide, tirzepatide that were approved by FDA for obesity treatment. For O-AOMs we considered bupropion, canagliflozin, dapagliflozin w/wo saxagliptin, empagliflozin w/wo linagliptin, lisdexamfetamine, metformin w/wo (liflozin, liptin, litazone,), topiramate, zonisamide. Specifically, exposure to an O-AOM ingredient for less than 30 days without a neighboring (30 days before or after) record was removed to allow for occasional use of O-AOMs for other purposes. While most of the O-AOMs are for diabetes treatment, we make no explicit distinction when the treatment course is longer than 30 days, in which case an impact on body weight should be anticipated. For the UT-Physician cohort, we searched the RxNav database ^32^ for all aliases of each drug ingredient and then searched the OHDSI Athena ^33^ database for concept ids of the original ingredient and all aliases. The concept ids were combined to represent a single corresponding ingredient. For COSMOS data, AOM usage records were identified by regular expression match of drug ingredient names.

#### Drug exposure period

For the UT-Physician cohort, data documented in the pharmacy medication dispense table was used as a proxy for drug exposure information. Specifically, the external drug dispense instant datetime was used as the exposure start date, and the start date plus the days supply was used as the exposure end date.

For the COSMOS cohort, the dispenses from EPIC pharmacy products as well as dispenses reported to Cosmos contributors via third-party interfaces were used as proxy for drug exposure information. The medication fill completion date was used as the exposure start date, and the exposure end date is defined as the start date plus the days’ supply. Both data elements are available in the MedicationDispenseFact table.

#### Drug dose

Drug dose was extracted for the UT-Physician cohort from the descriptive text associated with each dispensing record. After standardizing all dose units to milligrams, the daily dose was obtained by multiplying the dose amount with the dispensation unit count and then dividing by the days supplied. Dose was not analyzed for the COSMOS cohort due to only minimum dose was documented when dose is a range.

#### Treatment session

To effectively manage the complexity of AOM treatment in real-world scenarios, we meticulously designed algorithms to define a treatment session. This approach captures a continuous period during which the patient is exposed to generally the same active ingredient(s). Specifically, compared to an existing exposure record *REC_A_*, a new exposure record *REC_B_* belongs to the same treatment session (as *REC_A_*) if it 1) has exactly the same ingredient(s) as *REC_A_* and there is ≤40 days gap between start of *REC_B_* and end of *REC_A_*, or 2) has less ingredient than *REC_A_*, and has both exposure length and gap to *REC_A_* ≤40 days, or 3) has new ingredient(s) added within 40 days of initiating *REC_A_* and has exposure length ≤40 days. For all other circumstances, we consider *REC_B_* as a new treatment session.

#### Measurements

We retrieved measurements for body weight, body mass index (BMI), high-density lipoprotein (HDL), low-density lipoprotein (LDL), triglycerides, systolic blood pressure (SBP), diastolic blood pressure (DBP), and HbA1C level throughout each patient’s EHR history. Specifically, as an internal consistency check, body weight and BMI were analyzed independently rather than inferred from each other due to missing or discordant measurements in approximately 1% of records. For multiple measurements on a single day, the median value is used if the largest value is less than 20% higher than the minimum value or otherwise set to unavailable. For each treatment session, the ideal scenario is measurements are available on exactly both the start and end dates, which however is not always possible. To compensate for this, we considered four scenarios (Supp Table 3). Each treatment session belongs exclusively to one scenario with precedence decreasing from scenario 1 to 4. For each scenario we enforce a minimum of 30 days difference between two measurements to allow for meaningful treatment effect. Furthermore, with slight modification to previous report ^34^, we have adopted the equation (90 * exposure days / 365 days) to calculate for different drug exposure lengthens a customized allowable flanking range, which was further hard-clamped to between 30 days and 180 days to avoid overly stringent or loose requirements. Importantly, a measurement is disqualified if there are other AOM exposures between the measurement date and the date of exposure to the AOM of interest.

#### Calculation of weight change after AOM discontinuation

For each exposure record, the reference weight was determined by using the median body weight recorded from 30 days before to 30 days after the exposure end date. This reference weight was then compared with the median weights from six non-overlapping periods-30 to 90 days, 91 to 180 days, 181 days to 1 year, 1 to 1.5 years, 1.5 to 2 years, and over 2 years-to calculate the percentage changes in weight. Additionally, for a weight measurement after exposure to be considered valid, there must be no exposure to any AOMs from the end of the exposure period until the weight measurement is taken.

## Statistical analysis

Continuous data are summarized as medians and interquartile ranges (IQR). Categorical data are presented as frequencies and percentages. For percentage changes of measurements before and after treatment, the values beyond outer fence (Q3 + 3*IQR, Q1 - 3*IQR) were removed.

## Results

### Baseline characteristics of the study cohorts

The baseline characteristics of the two study cohorts, including those exposed to AOMs, are shown in Table 1. AOM exposure was recorded in 11.7% of the UT-Physician cohort and 27.9% of the COSMOS cohort, reflecting differences in observation periods (2020-2025 vs. 2018-2024) and population coverage (regional Houston-based vs. nationwide). In the UT-Physician cohort, AOM use increased markedly after mid-2020 (Supp Figure 1), a trend similarly observed in COSMOS.

**Table 1.** Baseline characteristics of the study cohorts

|  | UT-Physician (Jan.2015-Jun.2025) |  |  | COSMOS (Jan.2018-Dec.2024) |  |  |
| --- | --- | --- | --- | --- | --- | --- |
|  | Total Cohort | AOMs exposure <sup>c</sup> | Exposure % | Total Cohort <sup>d</sup> | AOMs exposure <sup>c</sup> | Exposure % |
| **Count** | 610,372 | 71,318 | 11.7% | 3,637,799 | 1,013,222 | 27.9% |
| **Gender** |  |  |  |  |  |  |
| Male | 236,225(39%) | 26,411(37%) | 11% | 1,506,126(41%) | 359,409(35%) | 24% |
| Female | 345,754(57%) | 44,890(63%) | 13% | 2,130,928(59%) | 653,616(65%) | 31% |
| **Race** |  |  |  |  |  |  |
| White | 219,245(36%) | 29,176(41%) | 13% | 2,431,904(67%) | 699,345(69%) | 29% |
| Black or AA | 139,401(23%) | 16,202(23%) | 12% | 616,602(17%) | 183,704(18%) | 30% |
| Asian | 16,246(3%) | 2,955(4%) | 18% | 79,404(2%) | 19,691(2%) | 25% |
| Others | 2,267(0.4%) | 310(0.4%) | 14% | 210,510(5.8%) | 47,133(4.7%) | 22% |
| Unknown | 233,212(38%) | 22,673(32%) | 10% | 299,379(8%) | 63,349(6%) | 21% |
| **Age at first diagnosis <sup>a</sup>** | 51(38, 65) | 54(43, 67) | - | 48(34, 63) | 50(38, 62) | - |
| **Diagnosis category <sup>b</sup>** |  |  |  |  |  |  |
| Obesity | 237,560(39%) | 26,925(38%) | 11% | 2,248,681(62%) | 626,210(62%) | 28% |
| Morbid obesity | 107,197(18%) | 13,957(20%) | 13% | 755,373(21%) | 264,330(26%) | 35% |
| Localized adiposity | 2,110(0.3%) | 272(0.4%) | 13% | 10,094(0.3%) | 2,892(0.3%) | 29% |
| Extreme obesity | 2,042(0.3%) | 172(0.2%) | 8% | 8,503(0.2%) | 2,590(0.3%) | 30% |
| Drug-induced obesity | 664(0.1%) | 197(0.3%) | 30% | 4,487(0.1%) | 1,430(0.1%) | 32% |
| Overweight | 260,771(43%) | 29,789(42%) | 11% | 610,661(17%) | 115,770(11%) | 19% |

<sup>a</sup> Values shown as median(1st quartile, 3rd quartile)
<sup>b</sup> For patients with multiple different obesity diagnoses, the first diagnosis is used. Simple obesity merged into obesity
<sup>c</sup> Based on pharmacy dispensing record available since 2020/05/09 for UT-Physician and 2007/04/07 for COSMOS, respectively
<sup>d</sup> 1/10 of the patients were randomly selected for final analysis

### AOMs exposure in the UT-Physician cohort

Analysis of prescription data indicates F-AOMs were rarely prescribed before 2015 (Supp Figure 1a). From 2015 to 2020, the F-AOMs prescription were dominated by liraglutide, phentermine, and bupropion naltrexone. Significant rise in prescription (Supp Figure 1a, 1b) and dispensing (Supp Figure 1c, 1d) of both F-AOMs and O-AOMs was observed after 2020, mainly attributable to semaglutide and metformin series. Tirzepatide stepped into the market after mid-2022, and became as popular as semaglutide after mid-2024.

Between 2020 and 2025, the most prevalent F-AOMs were semaglutide (29%), tirzepatide (13%), and phentermine (8%) among patients with AOM exposure. The leading O-AOMs were metformin (68%), bupropion (16%), and empagliflozin (14%). Combination use was also common, particularly metformin-semaglutide (11%) and metformin-tirzepatide (4%). Across all AOMs, short-term exposure (≤112 days) represents the predominant exposure type (Supp Figure 3).

Among patients with AOM exposure, 48% received a single AOM, 20% received two AOMs, and 32% were exposed to three or more AOMs; in extreme cases, 15 patients had exposure to ten or more AOMs. Focusing on F-AOMs, 27% of patients received one F-AOM, 8% received two, and 2% received three or more.

### Number and length of treatment sessions

We define a treatment session as a period of continuous exposure to a given drug ingredient or combination of ingredients. Transitioning to a new treatment session often entails physical, financial, and psychological burdens for patients and typically reflects inadequate efficacy or suboptimal response to the preceding session. Overall, [U:28%; C:33%] of patients documented only a single treatment session during the corresponding observation period, whereas the remaining patients experienced two or more sessions. Subgroup analyses further indicated that, across both cohorts, patients with overweight generally had fewer treatment sessions, while those with morbid obesity had more treatment sessions than other diagnostic categories (Figure 2).

**Figure 2.**
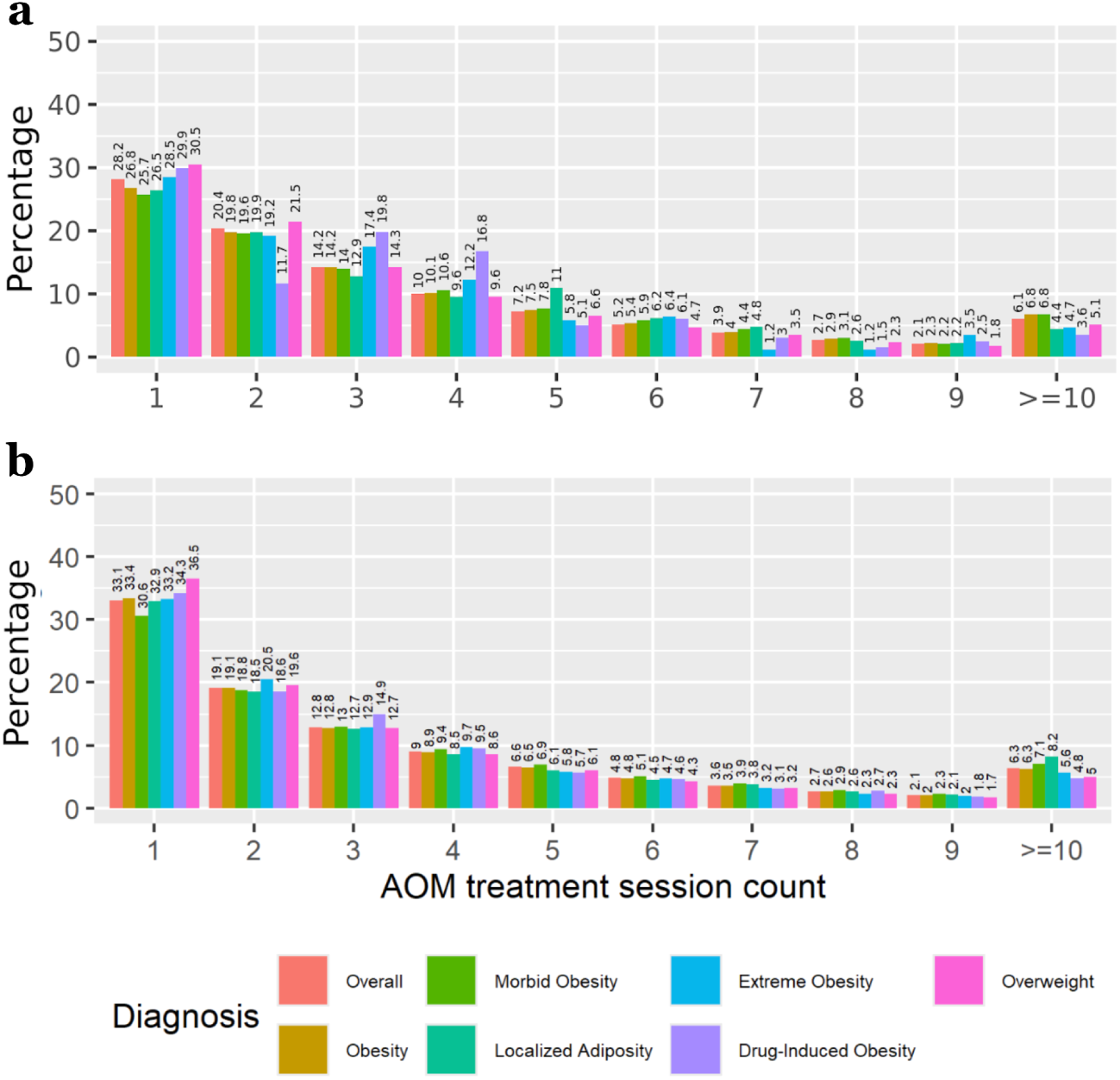
Distribution of treatment session counts in patients with different diagnoses in the (a) UT-Physician and (b) COSMOS cohort.

**Figure 3.**
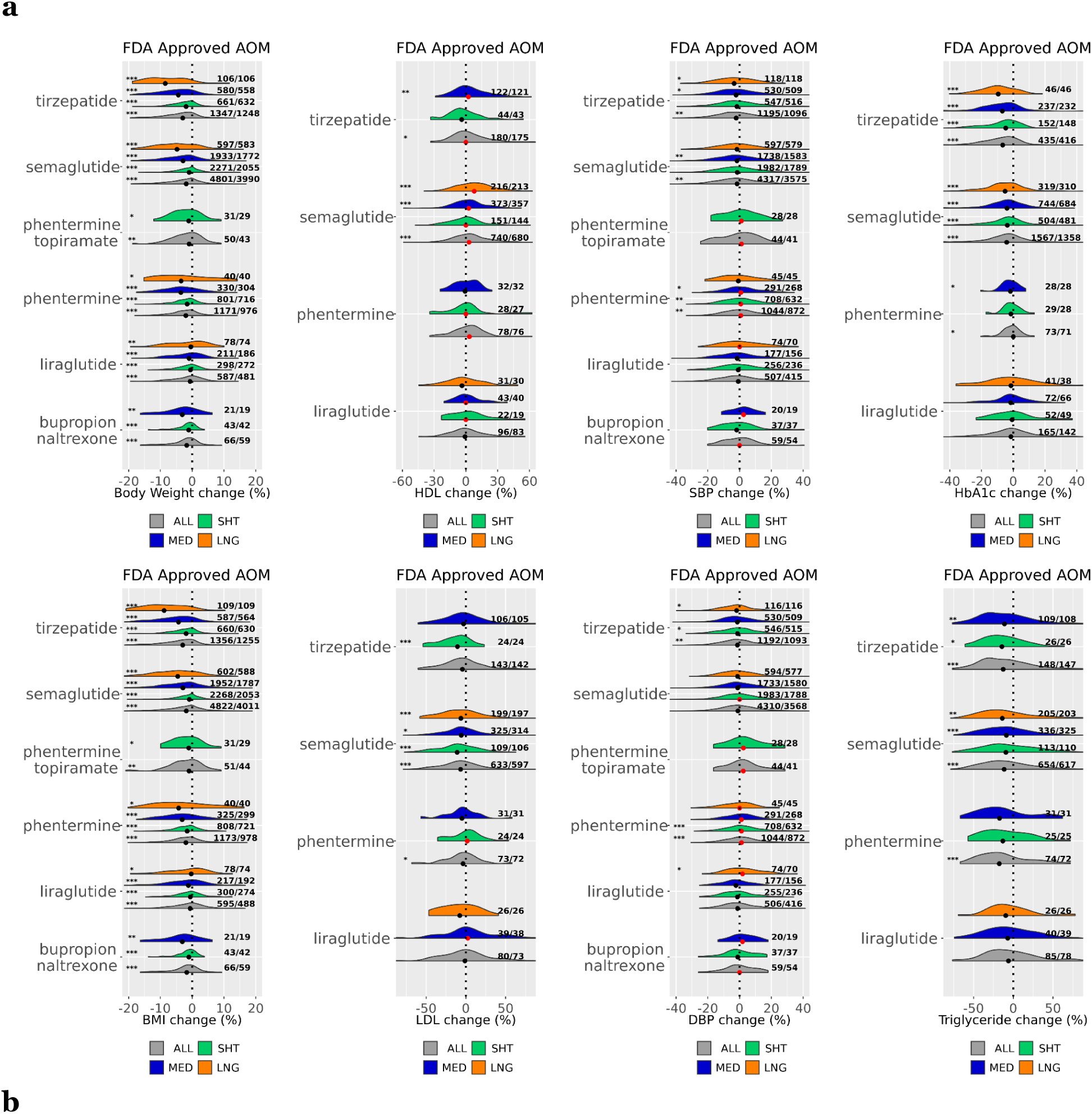
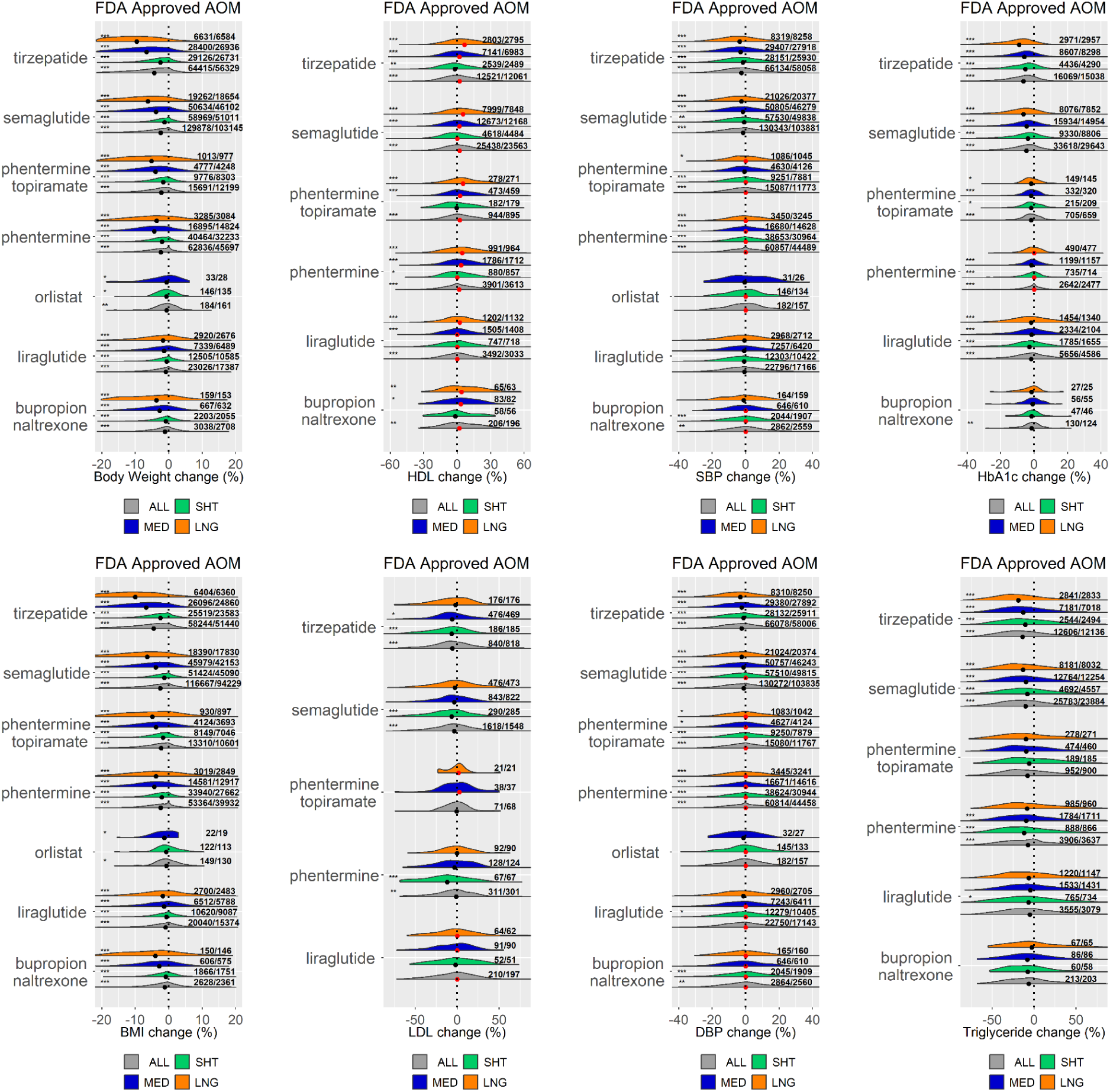
Distribution of percentage changes in obesity relevant metabolic metrics after various lengths (SHT: Short-term; MED: Medium-term; LNG: Long-term) of F-AOM exposure for a) UT-Physician cohort, and b) COSMOS cohort. Result displayed for each drug and category with a minimum of 20 observations. Black and red dots depict median values <0 and ≥0, respectively. Numbers on the right of each distribution shown as (number of observed records/number of unique patients). The asterisks on the left denote significance (p≤0.05 (*), p≤0.01 (**), and p≤0.001(***)) of two-tailed z-test against the null hypothesis of mean value 0.

The median treatment session lengths were [U:2.7; C:2.7] months and [U:3.0; C:3.1] months for F-AOMs and O-AOMs, respectively. For semaglutide, the median lengths were [U:2.8: C:3.0] months, with maximum session lengths of [U:65; C:87] months. For tirzepatide, the median lengths were [U:3.0; C:3.5] months, with maximum session lengths of [U:36, C:59] months being observed as of this writing. The distribution of treatment session lengths for each AOMs in UT-Physician is shown in Supp Figure 2.

### Responses of obesity relevant metabolic metrics to AOMs

For both cohorts, we investigated the effect of short to long term exposure (short-term: 30 to 112 days; medium-term: 113 to 365 days; long-term: >365 days) ^34^ to F-AOMs (Figure 3) and O-AOMs (Supp Figure 4). The metabolic metrics that are relevant to obesity, including body weight, BMI, HDL, LDL, SBP, DBP, HbA1c, and triglyceride levels were analyzed on their percentage changes before and after AOM exposure. In the following report, we employ both visual judgment of distributions and statistical significance tests to 1) prevent overlooking large effect sizes in exposures with small sample sizes, and 2) avoid misinterpreting high statistical significance purely due to large sample sizes but with less meaningful effect sizes. We summarize the major observations as follows.

**Figure 4.**
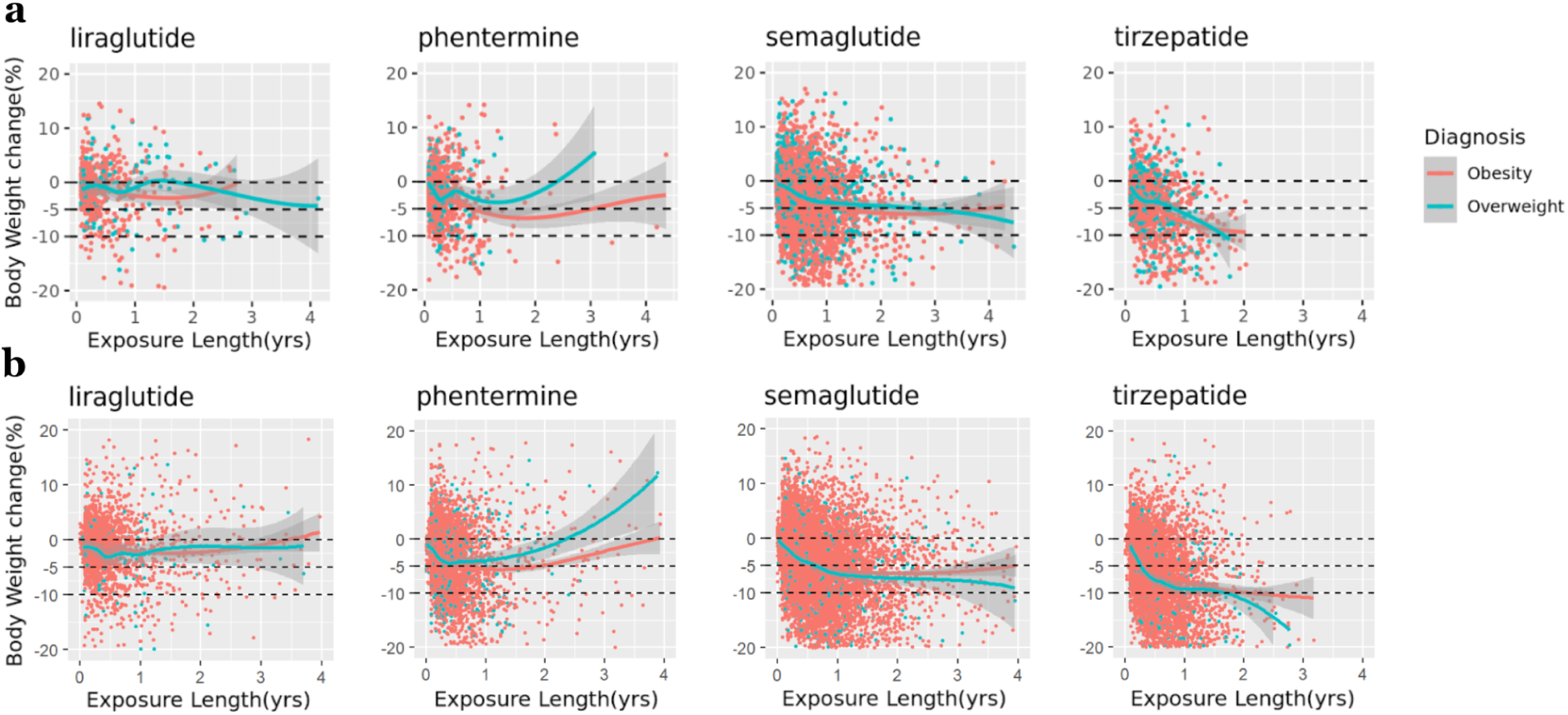
Correlation between exposure durations of top four F-AOMs and percentage changes in body weight in a) UT-Physician cohort, and b) COSMOS cohort, categorized by type of diagnosis (all obesities vs overweight), with LOESS smooth lines applied using local regression.

### Response to F-AOMs (Figure 3) Body weight and BMI

Both tirzepatide and semaglutide were associated with substantial body weight loss, generally outperforming other F-AOMs at medium to long-term exposure durations. Weight loss magnitude increased with longer exposure and reached strong statistical significance (p < 0.001), with clearly discernible effect sizes observed in both cohorts. In contrast, under short-term exposure, semaglutide demonstrated comparable effectiveness to the phentermine-topiramate combination, which conferred greater weight reduction than phentermine monotherapy on long-term exposure. Across both cohorts, liraglutide was associated with only modest weight loss across all exposure-length subgroups. Analyzing BMI independently yields results consistent with those observed for body weight, despite minor differences in patient sample sizes and exposure counts.

### Lipid levels

All F-AOMs were associated with some degree of triglyceride reduction, with tirzepatide, semaglutide, and phentermine/topiramate demonstrating the most pronounced benefits across all exposure durations. In contrast, consistent reductions in LDL cholesterol were observed only for tirzepatide and semaglutide during short- to medium-term exposure, with these effects attenuating at longer exposure durations. Conversely, improvements in HDL cholesterol levels were primarily evident with long-term exposure across all F-AOMs.

### Blood pressure and HbA1c levels

Across both cohorts, only tirzepatide and semaglutide were associated with significant reductions in HbA1c levels, with larger effect size on longer exposure duration. Both AOMs also demonstrated modest reductions in SBP and DBP, with effect sizes generally increasing with longer exposure. Liraglutide exhibited only minimal HbA1c-lowering effects and no consistent impact on blood pressure. In contrast, other F-AOMs showed no appreciable effects on either HbA1c or blood pressure.

### Response to O-AOMs (Supp Figure 4,5) Body weight and BMI

Among O-AOMs, only long-term exposure to metformin-based regimens and SGLT2 inhibitors (empagliflozin, dapagliflozin, and canagliflozin) was associated with modest body weight reduction across both cohorts, with effect sizes substantially smaller than those observed for tirzepatide and semaglutide. Most other O-AOMs did not demonstrate meaningful weight-loss benefits in either cohort. Notably, long-term exposure to lisdexamfetamine was consistently associated with weight gain across both cohorts and reached statistical significance (p < 0.001) in the COSMOS cohort.

Combining metformin with tirzepatide or semaglutide did not confer additional weight-loss benefits compared with tirzepatide or semaglutide monotherapy. BMI-based analyses yielded findings consistent with those observed for body weight.

### Lipid levels

All O-AOMs demonstrated some degree of triglyceride reduction across both cohorts. Notably, the magnitude of this effect was consistently greater with short-term exposure than with medium- or long-term exposure. A similar pattern was observed for LDL, with most O-AOMs (except zonisamide and lisdexamfetamine) showing mild LDL reductions predominantly during short-term use. In contrast, effects on HDL were modest and inconsistent; when present, HDL increases were largely seen on long-term exposures.

### Blood pressure and HbA1c levels

None of the O-AOMs showed a meaningful effect on either SBP or DBP. The metformin series and SGLT2 inhibitors modestly reduced HbA1c, primarily with short-term exposure, with effect sizes substantially smaller than those observed for F-AOMs.

### Variations in body weight responses to AOMs

While several AOMs demonstrated statistically significant weight-loss effects at the population level, individual responses varied substantially. For example, long-term exposure to metformin was associated with a modest but significant reduction in body weight (median ∼1.5%, *p*≤0.001; Supp Figure 5); however, [U:74%; C:72%] of patients failed to achieve ≥5% weight loss, and [U:40%; C:39%] experienced weight gain (Supp Tables 1 and 2).

**Figure 5.**
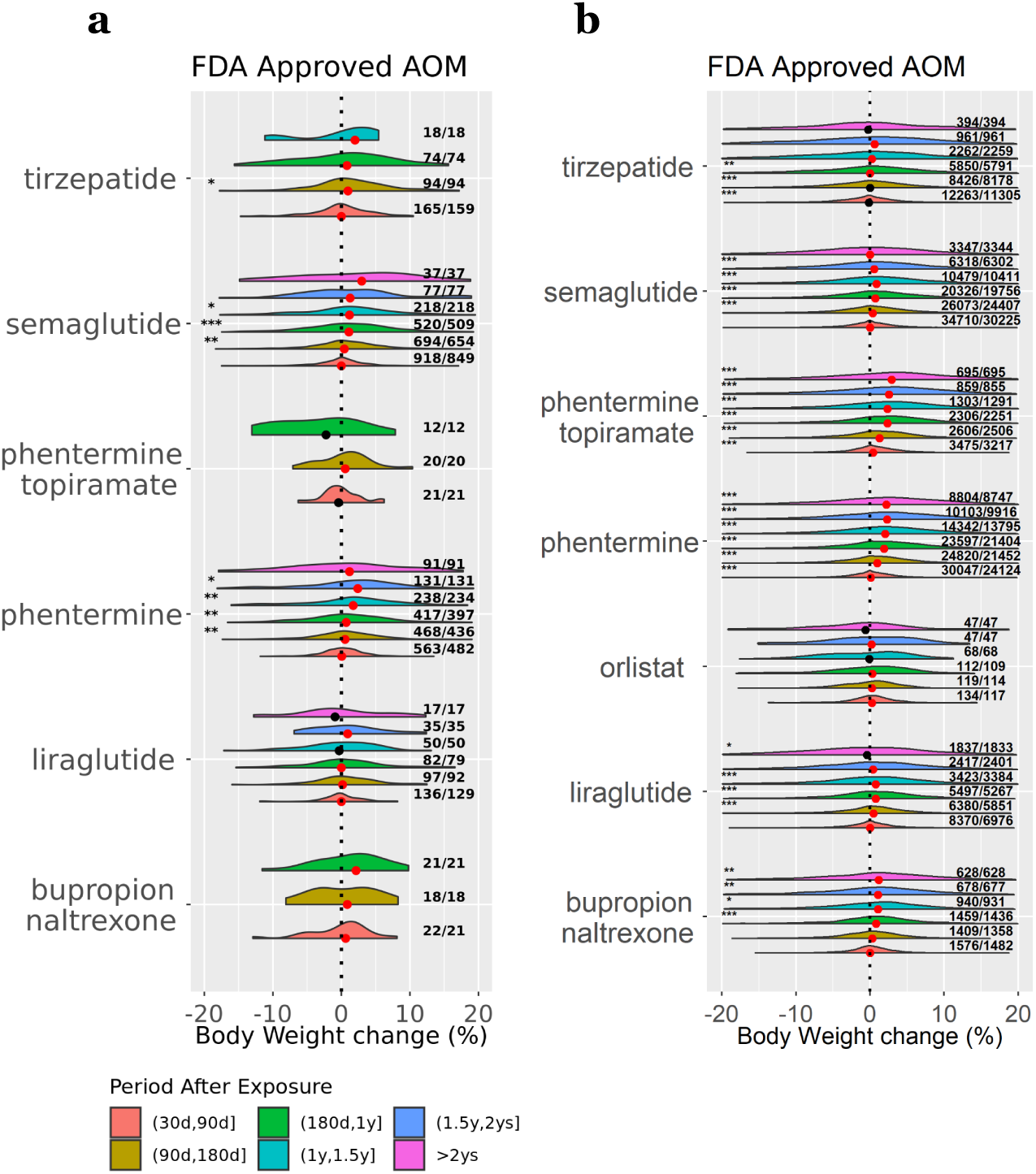
Body weight percentage changes after discontinuing F-AOMs, using body weight at the end of exposure as reference weight. a) UT-Physician cohort. b) COSMOS cohort.

**Table 2.** Response of body weight to F-AOMs

| Ingredient<sup>a</sup> | Median Dose | UT-Physician |  |  |  |  |  | COSMOS |  |  |  |  |
| --- | --- | --- | --- | --- | --- | --- | --- | --- | --- | --- | --- | --- |
|  |  | Term | Exposure Length (wks) | Records/Patients | ≥10% Loss | ≥5% Loss | Weight Gain | Exposure Length (wks) | Records/Patients | ≥10% Loss | ≥5% Loss | Weight Gain |
| bupropion<br>naltrexone | 360mg/d | Short | 7[4,8]<sup>b</sup> | 43/42 | <5<sup>d</sup> | 6(14%) | 14(33%) | 7[4,8] | 2203/2055 | 18(1%) | 248(11%) | 834(38%) |
|  |  | Medium | 28[22,30] | 21/19 | <5 | 7(33%) | 6(29%) | 26[20,28] | 667/632 | 88(13%) | 246(37%) | 202(30%) |
|  |  | Long | 80[70,80] | <11<sup>c</sup> | - | - | - | 70[59,84] | 159/153 | 46(29%) | 75(47%) | 49(31%) |
| liraglutide | 3mg/d | Short | 9[6,9] | 298/272 | 7(2%) | 39(13%) | 124(42%) | 8[5,9] | 12505/10585 | 244(2%) | 1494(12%) | 5234(42%) |
|  |  | Medium | 27[21,29] | 211/186 | 15(7%) | 59(28%) | 89(42%) | 27[21,29] | 7339/6489 | 612(8%) | 1922(26%) | 2906(40%) |
|  |  | Long | 78[62,84] | 78/74 | 8(10%) | 25(32%) | 36(46%) | 78[62,94] | 2920/2676 | 389(13%) | 902(31%) | 1178(40%) |
| phentermine | 37.5mg/d | Short | 8[4,8] | 801/716 | 14(2%) | 130(16%) | 247(31%) | 8[4,8] | 40464/32233 | 1521(4%) | 8997(22%) | 11251(28%) |
|  |  | Medium | 26[19,28] | 330/304 | 46(14%) | 125(38%) | 97(29%) | 25[20,28] | 16895/14824 | 3231(19%) | 7756(46%) | 3974(24%) |
|  |  | Long | 67[57,85] | 40/40 | 10(25%) | 18(45%) | 14(35%) | 71[59,87] | 3285/3084 | 777(24%) | 1444(44%) | 1115(34%) |
| phentermine<br>topiramate | 7.5(top)-46<br>(phe)<br>mg/d | Short | 8[5,9] | 31/29 | <5 | 8(26%) | 10(32%) | 8[5,8] | 9776/8303 | 315(3%) | 1900(19%) | 3016(31%) |
|  |  | Medium | 19[17,21] | 13/12 | <5 | <5 | 7(54%) | 26[20,28] | 4777/4248 | 903(19%) | 2077(43%) | 1242(26%) |
|  |  | Long | 67[60,85] | <11 | - | - | - | 71[59,82] | 1013/977 | 309(31%) | 513(51%) | 287(28%) |
| semaglutide | 1.7mg/w<br>(Injection)<br>7mg/d(Tab<br>let) | Short | 9[7,10] | 2271/2055 | 39(2%) | 325(14%) | 887(39%) | 9[6,9] | 58969/51011 | 1624(3%) | 9477(16%) | 20276(34%) |
|  |  | Medium | 27[21,29] | 1933/1772 | 264(14%) | 704(36%) | 541(28%) | 28[22,30] | 50634/46102 | 9736(19%) | 21679(43%) | 12743(25%) |
|  |  | Long | 70[60,80] | 597/583 | 140(23%) | 291(49%) | 163(27%) | 72[60,82] | 19262/18654 | 6300(33%) | 10772(56%) | 3978(21%) |
| tirzepatide | 5mg/w | Short | 9[7,10] | 661/632 | 24(4%) | 151(23%) | 199(30%) | 9[6,9] | 29126/26731 | 1937(7%) | 8152(28%) | 7486(26%) |
|  |  | Medium | 27[21,29] | 580/558 | 114(20%) | 266(46%) | 121(21%) | 28[22,30] | 28400/26936 | 9335(33%) | 16701(59%) | 4505(16%) |
|  |  | Long | 65[58,69] | 106/106 | 45(42%) | 69(65%) | 8(8%) | 66[57,72] | 6631/6584 | 3179(48%) | 4700(71%) | 775(12%) |

<sup>a</sup> Orlistat not displayed due to insufficient number of observations for both cohorts<sup>b</sup> Value shown as median[IQR]<sup>c</sup> Statistics only calculated for 11 or more observed records<sup>d</sup> Records with count <5 masked for patient privacy

Long-term exposure to tirzepatide, the most effective F-AOM, resulted in a substantial median body-weight reduction of ∼9% (*p*≤0.001; Figure 3). In this group, [U:42%; C:48%] of patients achieved ≥10% weight loss, whereas [U:35%; C:29%] failed to achieve ≥5% weight loss, and [U:8%; C:12%] experienced weight gain by the end of exposure (Table 2). In comparison, long-term exposure to semaglutide was associated with weight gain in [U:27%; C:21%] of cases (Table 2).

Among F-AOMs, excluding orlistat due to limited sample size, liraglutide showed the poorest performance, resulting in weight gain in ≥40% of cases (Table 2). Among O-AOMs, bupropion, lisdexamfetamine, and zonisamide exhibited the least favorable outcomes, with weight gain observed in ≥50% of cases on various exposure durations across both cohorts (Supp Tables 1 and 2).

Scatterplots of individual F-AOM exposure records showed that, for liraglutide and phentermine, the association between exposure duration and body weight change was generally weak beyond six months (Figure 4). For semaglutide, longer exposure was associated with greater weight loss up to approximately one year, after which no additional population-level benefit was observed. Tirzepatide demonstrated the most rapid weight reduction within the first year of treatment; beyond one year, weight-loss effects were more pronounced among patients with overweight diagnosis. Across all F-AOMs, substantial interindividual variability in weight-loss response was observed, largely independent of exposure duration. Additional subgroup analyses revealed comparable degrees of variability across sex, race, diagnosis category, and age at treatment initiation (data not shown).

### Weight changes after AOMs discontinuation

We evaluated body-weight changes across six discrete, non-overlapping periods following discontinuation of F-AOMs (Figure 5) and O-AOMs (Supp Figures 6,7).

Although individual responses to AOM discontinuation varied substantially, several consistent patterns emerged. Among F-AOMs, phentermine-topiramate, phentermine, and bupropion-naltrexone demonstrated the most pronounced and consistent trends toward gradual weight regain in both cohorts after treatment cessation. In contrast, tirzepatide and semaglutide were associated with only modest weight gain up to 1.5 years post-discontinuation. Notably, when co-administered with metformin, both tirzepatide and semaglutide exhibited sustained weight-loss effects after discontinuation in the COSMOS cohort (Supp Figure 7). Among O-AOMs, lisdexamfetamine, zonisamide, and topiramate were associated with substantial weight regain across multiple post-exposure periods in both cohorts, whereas discontinuation of SGLT2 inhibitors and metformin generally did not result in significant weight regain.

## Discussion

With two data sources managed under different data model systems-the OMOP Common Data Model and the COSMOS Caboodle Data Model-and patient cohorts characterized by highly diversified population demographics, we achieved notably consistent insights regarding responses of obesity relevant metabolic metrics to AOMs and body weight changes after AOMs cessation. Overall, our findings highlight the effectiveness of F-AOMs, particularly GLP-1/GIP receptor agonists, in weight loss, glycemic control, and improving lipid profiles in a limited proportion of patients. Our results further underscore secondary use of EHR as a valuable strategy for retrospective verification of treatment responses in real-world scenarios, aligning with the increasing popularity of using EHR in RCTs ^35^ and to derive real-world evidence ^36^. The consistency across data models underscores the potential of EHR to support comparative analyses and improve treatment evaluations across varied clinical questions ^37^.

In this real-world evidence based study, several F-AOMs demonstrated substantially smaller weight-loss effects than those reported in RCTs ^23^. For example, semaglutide was reported to achieve 10% weight loss target in at least 69.1% of participants after 68 weeks at 2.4mg/week in an RCT ^27^, whereas we observed that no more than 33% of patients reached this threshold after a median of 72 weeks of exposure. Similarly, in another RCT ^28^, tirzepatide achieved 10% weight loss target in at least 68.5% of patients after 72 weeks exposure at 5mg per week, the lowest dose group in the trial. In contrast, only 48-50% of patients achieved the 10% target after a median of 72 weeks exposure in our additional analysis. However, it is noteworthy that our tirzepatide results align more closely with an RCT conducted on patients with type 2 diabetes ^38^. In this trial, the 5mg group achieved 5% weight loss in 34% patients and 10% weight loss in 65% patients after 40 weeks. The results closely match our corresponding results of 32-41% and 61-65% after a median of 40 weeks exposure.

The disparity can be explained as follows. First, dosing in real-world clinical practice tends to be more conservative for certain AOMs. For example, the median dose of injectable semaglutide observed in the UT-Physician cohort was 1.7 mg/week, substantially lower than the 2.4 mg/week maintenance dose in most RCTs. In addition, we observed that approximately 8% of semaglutide were administered as oral formulations, which has generally lower efficacy than the injectable form. Secondly, the rigorous inclusion/exclusion criteria of RCTs may also play a crucial role. The observation that tirzepatide performance in our study aligns better with the trial involving diabetes patients ^38^ than without diabetes ^28^ may reflect the confounding effect of comorbidities in real-world practice. Indeed, in the UT-Physician cohort about half of the patients screened for overweight had at least one HbA1c measurement in the prediabetic range (5.7% to 6.5%), signifying the confounding effect of comorbidities. Thirdly, medication adherence in real-world practice is generally less strict than in RCTs. Adherence issues include non-fulfillment (never initiated), non-persistence (stop taking after initiation), non-conforming (skipping dose, incorrect does) ^39^ have been reported in various AOM adherence studies ^40,41,42^, and likely diminishes AOM effectiveness. Finally, we do not rule out the potential bias stemming from pharmaceutical industry sponsorship in many of these RCTs. Overall, our results reflect the general population’s response to AOMs amidst these real-world factors.

Our findings of weight regain after stopping several F-AOMs and O-AOMs were consistently observed in both cohorts and aligned with previous research on tirzepatide ^43^, semaglutide ^44^, bupropion-naltrexone ^45^, phentermine ^34^, zonisamide ^46^, and topiramate ^47^. In addition, the most serious weight regain was observed after discontinuing lisdexamfetamine, which however was not formally documented elsewhere to the best of our knowledge. Interestingly, while metformin co-administration with tirzepatide or semaglutide was associated with smaller weight reductions than tirzepatide or semaglutide monotherapy, it was also associated with notably less weight regain following treatment discontinuation. Consistently, prior work suggests that metformin may attenuate weight regain after cessation of semaglutide ^48^, although comparable evidence for tirzepatide is currently lacking.

Substantial individual variations were observed regardless of the type of AOM and duration of exposure. For O-AOMs, variations could be due to the treatment not always being aimed at obesity management, so early response might not influence the decision to continue or discontinue the treatment. This could partially explain the fluctuations in body weight changes over short to long term exposures. Nevertheless, we didn’t specifically observe less variation in body weight responses to medium-term and long-term F-AOMs, which are supposedly based on short-term effectiveness. This suggests that some patients may not be promptly modifying their treatment plans in response to early outcomes, or may develop tolerance, or exhibit reduced adherence after a period of continued exposure.

Overall, the remarkably inconsistent AOM responses at individual level underscore recent move away from the oversimplified classification by BMI ^49^ toward more sophisticated staging strategies ^50^ and phenotyping-guided therapies ^51^. Recent review articles ^52,53^ highlighted the necessity of obesity deep phenotyping for precision medicine, as “every obese individual should be assessed based on their own specific and unique circumstances” ^54^. Our findings reinforce the significance of phenotyping-guided therapies to enhance both the effectiveness and durability of AOM therapy in routine clinical practice and advance precision obesity care.

## Conclusion

Our study confirmed the real-world effectiveness of AOMs, particularly F-AOMs, on a panel of obesity relevant metabolic metrics in a limited proportion of patients, highlighting substantial individual variability in treatment responses and notable gaps between EHR-based real-world evidence and clinical trial findings. These discrepancies underscore the challenges of generalizing results from clinical trials to the broader and more diverse populations encountered in routine clinical practice. The marked heterogeneity in individual responses to both FDA-approved and Off-label AOMs emphasizes the need for a personalized approach to obesity pharmacotherapy, which may enhance treatment effectiveness and improve long-term adherence, thereby supporting more sustainable obesity care strategies.

## Limitations

First, we used pharmacy dispensing records as a proxy for AOM uptake, which does not constitute definitive evidence of patient adherence to prescribed medication regimens. Although medication adherence cannot be directly observed, follow-up body weight/BMI measurements after an AOM dispense may serve as a proxy for continued engagement in weight management care and may indirectly reflect medication use. Second, UT-Physician drug dispensing records were consistently available only after the 2020 EPIC transition, which may lead to an underestimation of AOM exposure between 2015 and 2020. However, because the majority of AOM prescriptions occurred after 2020, and our analyses focused on AOM responses following documented exposure, the overall impact of this limitation is likely minimal. Third, we observed a decline in recorded AOM exposures toward the end of 2024, which may reflect delays in pharmacy dispensing data being integrated into the EPIC system. This lag could result in an overestimation of the effects of certain AOM exposures. Fourth, because the COSMOS database includes healthcare systems using the EPIC platform, it may contain overlapping patient records with the UT-Physician database, particularly following UT-Physician’s transition to EPIC in 2020. Nevertheless, as post-2020 UT-Physician records account for less than 1% of the COSMOS dataset, the impact of potential overlap on our findings is expected to be minimal. Finally, due to computational constraints on the COSMOS platform, our analyses were limited to data from 2018 to 2024. This restriction may bias estimates of the overall effectiveness of AOMs that were more commonly used before 2018. In addition, since this study focused primarily on the effectiveness of AOMs on obesity relevant metabolic metrics, the analyses of potential adverse effects were beyond the scope of the current work and will be addressed in a separate study.

## Data Availability

The data supporting this study are not publicly available due to restrictions related to patient privacy and confidentiality. Access to the data may be granted upon reasonable request and with appropriate institutional approvals.

## List of abbreviations

AOM: Anti-Obesity Medication
BMI: Body Mass Index
BP: Blood Pressure BS: Bariatric Surgery
DBP: Diastolic Blood Pressure
EHR: Electronic Health Record
F-AOM: FDA-approved AOM
FDA: Food and Drug Administration
HDL: High-Density Lipoprotein
IQR: Interquartile Range
LDL: Low-Density Lipoprotein
O-AOM: Off-label AOM
RCT: Randomized Clinical Trial
SBP: Systolic Blood Pressure
UT: University of Texas

## Declarations

### Ethics approval and consent to participate

The University of Texas Institutional Review Board (IRB) approved the study, and the Ethics Committee waived the need for written informed consent from participants.

### Consent for publication

Not applicable

### Availability of data and materials

The data supporting this study are not publicly available due to restrictions related to patient privacy and confidentiality. Access to the de-identified UT-Physician data may be granted upon reasonable request and with appropriate institutional approvals.

## Authors’ contributions

XR led the study design, conducted data analysis, and drafted the manuscript. RL carried out data analysis for the COSMOS cohort. LW and SL contributed to manuscript preparation. AW oversaw the harmonization of the UT-Physician EHR with the OMOP CDM. MS provided expert insights as an obesity clinician. HL contributed to the study design and coordinated the overall project.

## Acknowledgement

Not applicable

