## Supplementary Text and Figures for "Current status of anti-obesity medications and performance, an EHR based survey"

**Supp Text 1 Obesity subtypes and OMOP Concept codes**

Obesity: 433736; Morbid obesity: 434005; Localized adiposity: 438731; Extreme obesity with alveolar hypoventilation: 4100857; Drug-induced obesity: 4097996; Simple obesity: 4217557

**Supp Text 2 Comorbidities for determining overweight**

prediabetes, diabetes, hypertension, metabolic syndrome, obstructive sleep apnea, polycystic ovary syndrome, insulin resistance, hyperlipidemia, fatty liver, non-alcoholic steatohepatitis, coronary artery disease, cerebrovascular accident, stroke, peripheral vascular disease, congestive heart failure, colon/breast/renal/endometrial/liver cancer, osteoarthritis, or with HbA1c >= 5.7 and < 6.5

**Supp Text 3 ICD-9 and ICD-10 diagnosis codes for determining obesity phenotype in COSMOS database**

ICD-9: (‘278’, ‘278.0’, ‘278.00’, ‘278.01’, ‘278.02’, ‘278.03’, ‘278.1’, ‘278.2’, ‘278.3’, ‘278.4’, ‘278.8’)

ICD-10: (‘E66’, ‘E66.0’, ‘E66.01’, ‘E66.09’, ‘E66.1’, ‘E66.2’, ‘E66.3’, ‘E66.8’, ‘E66.9’)

**Supp Text 4 ICD-9 and ICD-10 procedure codes for determining obesity-related surgery in COSMOS database**

ICD-9 procedure codes ( ‘42.62’, ‘43’, ‘43.5’, ‘43.7’, ‘43.8’, ‘43.81’, ‘43.82’, ‘43.89’, ‘43.9’, ‘43.91’, ‘43.99’, ‘44.3’, ‘44.31’, ‘44.32’, ‘44.39’, ‘44.9’, ‘44.95’, ‘44.96’)

ICD-10 procedure codes ('0D11076', '0D110J6', '0D110K6', '0D110Z6', '0D11476', '0D114J6', '0D114K6', '0D114Z6', '0D11876', '0D118J6', '0D118K6', '0D118Z6', '0D12076', '0D120J6', '0D120K6', '0D120Z6', '0D12476', '0D124J6', '0D124K6', '0D124Z6', '0D12876', '0D128J6', '0D128K6', '0D128Z6', '0D13076', '0D130J6', '0D130K6', '0D130Z6', '0D13476', '0D134J6', '0D134K6', '0D134Z6', '0D13876', '0D138J6', '0D138K6', '0D138Z6', '0D15076', '0D150J6', '0D150K6', '0D150Z6', '0D15476', '0D154J6', '0D154K6', '0D154Z6', '0D15876', '0D158J6', '0D158K6', '0D158Z6', '0D16074', '0D16079', '0D1607B', '0D1607L', '0D160J4', '0D160J9', '0D160JB', '0D160JL', '0D160K4', '0D160K9', '0D160KB', '0D160KL', '0D160Z4', '0D160Z9', '0D160ZB', '0D160ZL', '0D163J4', '0D16474', '0D16479', '0D1647B', '0D1647L', '0D164J4', '0D164J9', '0D164JB', '0D164JL', '0D164K4', '0D164K9', '0D164KB', '0D164KL', '0D164Z4', '0D164Z9', '0D164ZB', '0D164ZL', '0D16874', '0D168J4', '0D168K4', '0D168Z4', '0DB40ZX', '0DB40ZZ', '0DB43ZX', '0DB43ZZ', '0DB44ZX', '0DB44ZZ', '0DB47ZX', '0DB47ZZ', '0DB48ZX', '0DB48ZZ', '0DB60Z3', '0DB60ZX', '0DB60ZZ', '0DB63Z3', '0DB63ZX', '0DB63ZZ', '0DB64Z3', '0DB64ZX', '0DB64ZZ', '0DB67Z3', '0DB67ZX', '0DB67ZZ', '0DB68Z3', '0DB68ZX', '0DB68ZZ', '0DB74ZX', '0DB74ZZ', '0DT40ZZ', '0DT44ZZ', '0DT47ZZ', '0DT48ZZ', '0DT60ZZ', '0DT64ZZ', '0DT67ZZ', '0DT68ZZ', '0DT74ZZ', '0F140D4', '0F140Z4', '0F144D4', '0F144Z4', '0F150D4', '0F150Z4', '0F154D4', '0F154Z4', '0F160D4', '0F160Z4', '0F164D4', '0F164Z4', '0F170D4', '0F170Z4', '0F174D4', '0F174Z4', '0F180D4', '0F180Z4', '0F184D4', '0F184Z4', '0F190D4', '0F190Z4', '0F194D4', '0F194Z4', '0F1D0D4', '0F1D0Z4', '0F1D4D4', '0F1D4Z4').

**a	b**

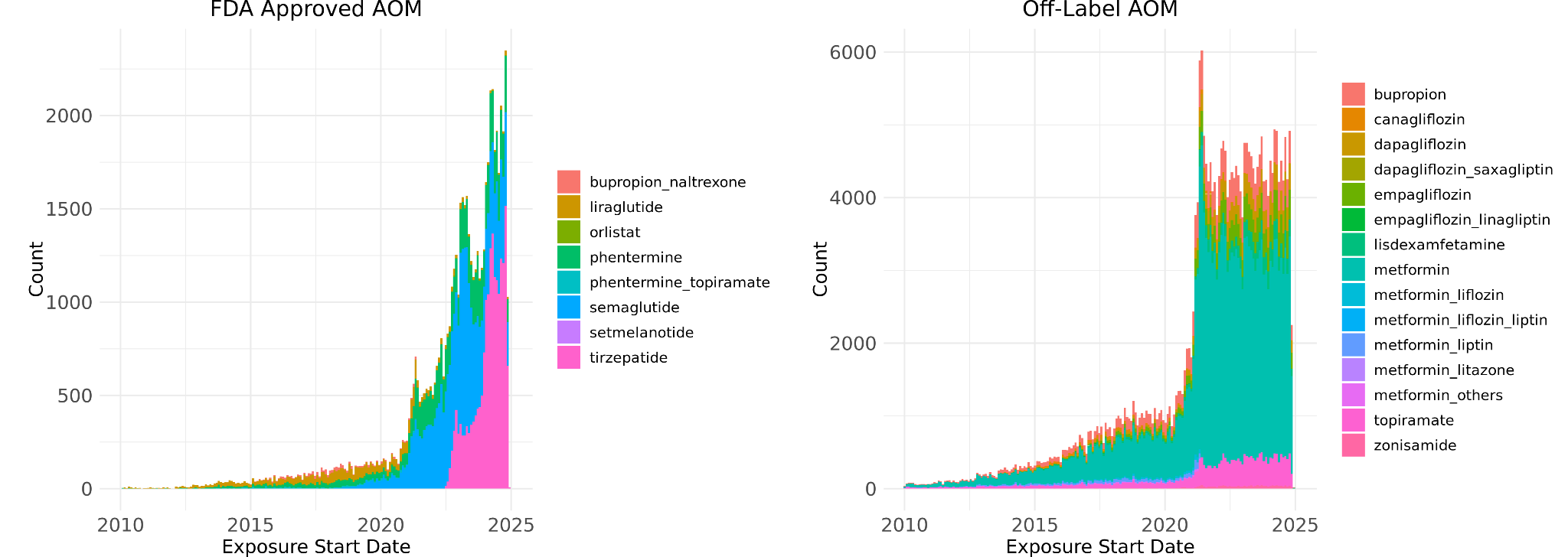

**c	d**

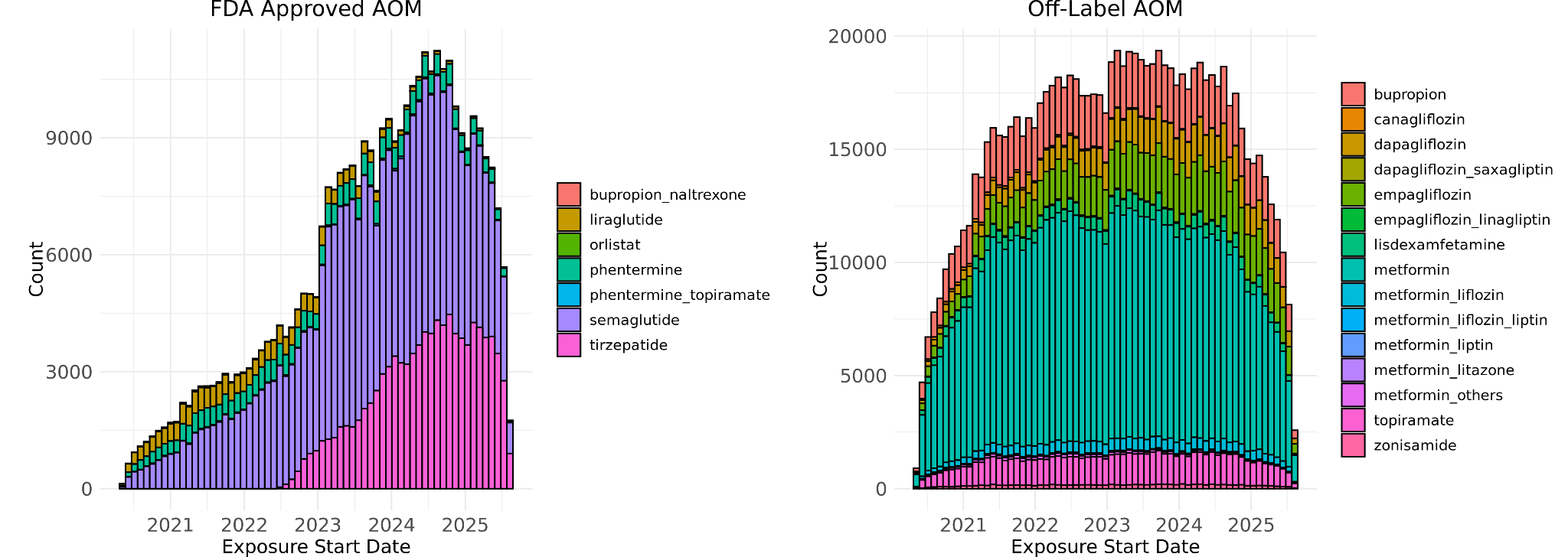

**Supp Figure 1** Stacked histograms of prescription records for (a) F-AOMs and (b) O-AOMs from 2010 to 2025, and dispensing records for (c) F-AOMs and (d) O-AOMs from May 2020 to September 2025 in the UT-Physician cohort.

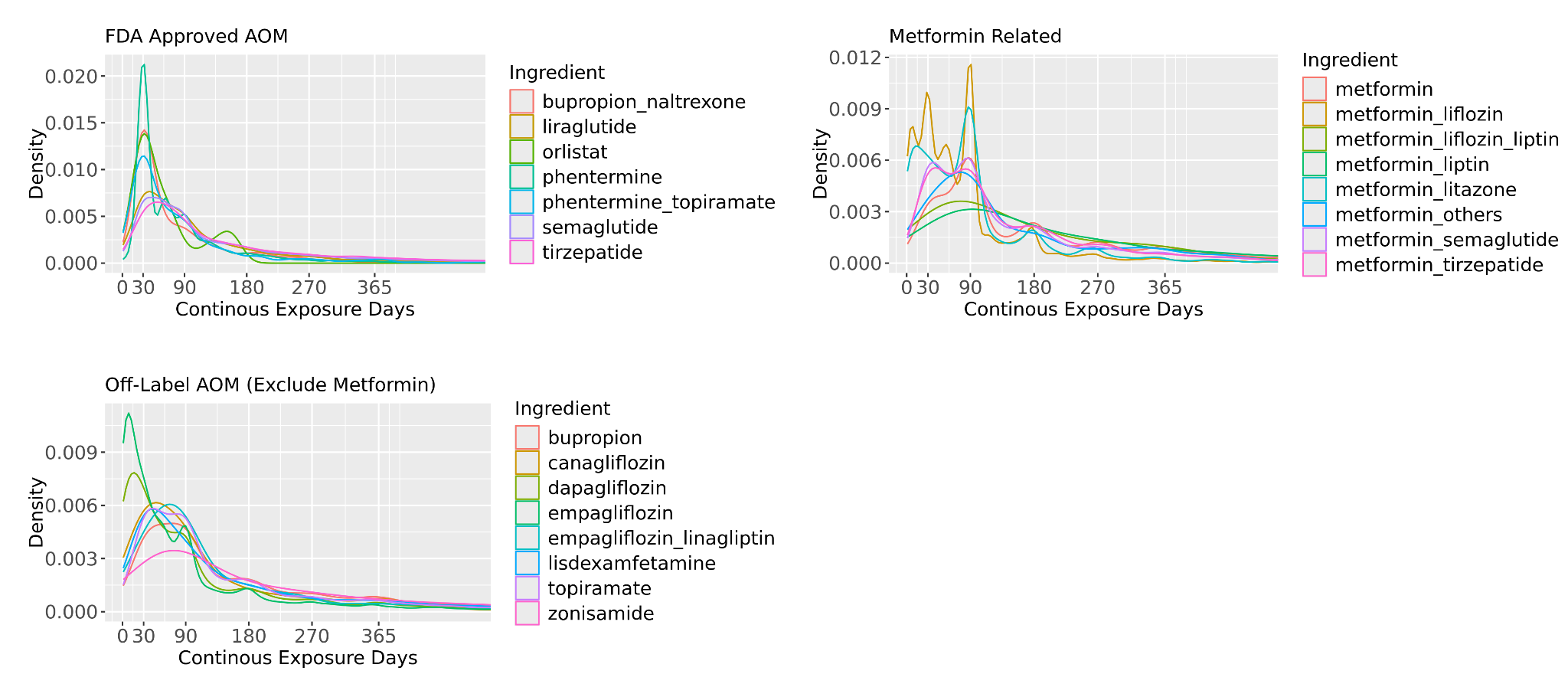

**Supp Figure 2** Distribution of treatment session length for F-AOM and O-AOM in the UT-Physician cohort

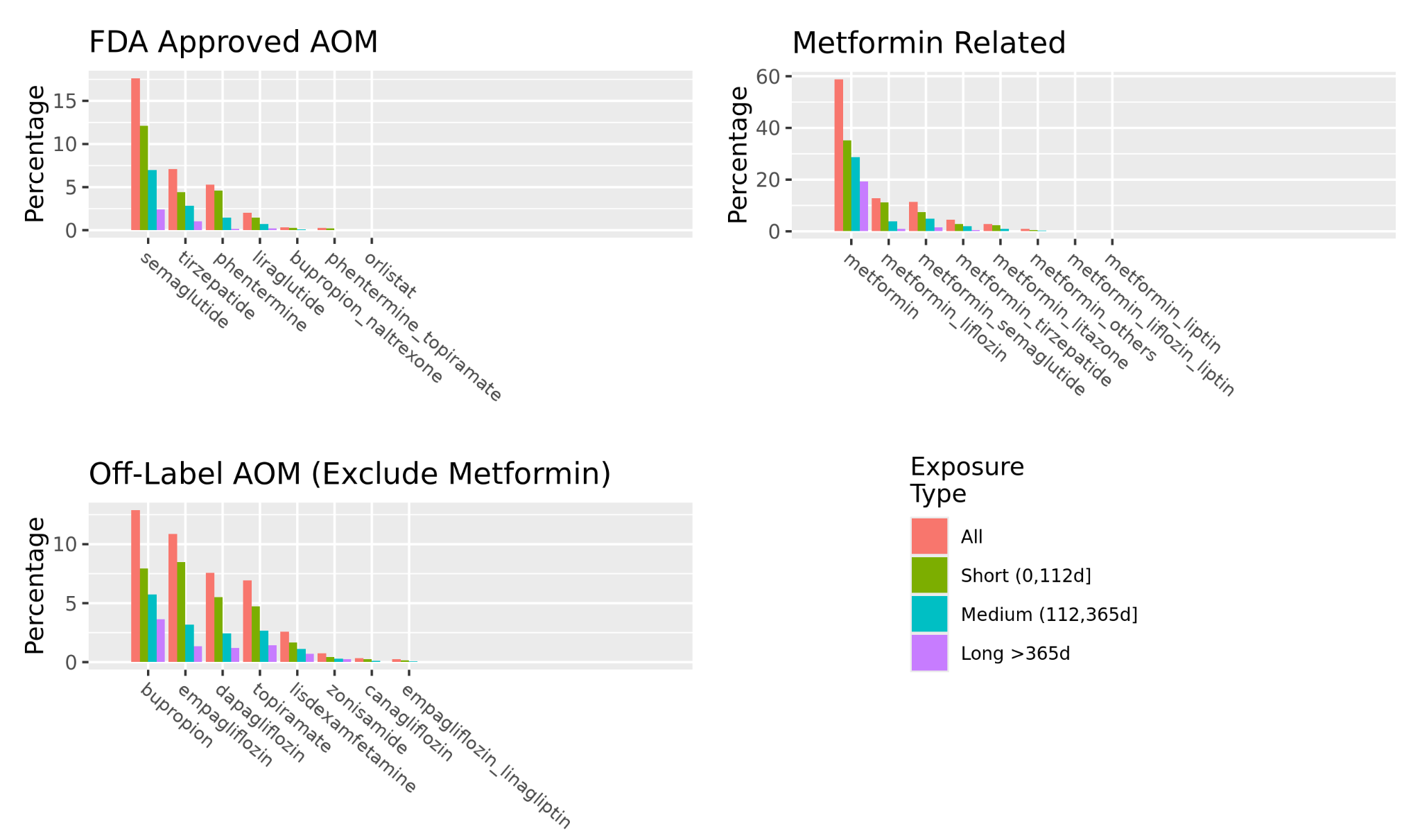

**Supp Figure 3** Prevalence of F-AOM and O-AOM among patient with AOM exposure in the UT-Physician cohort

**a**

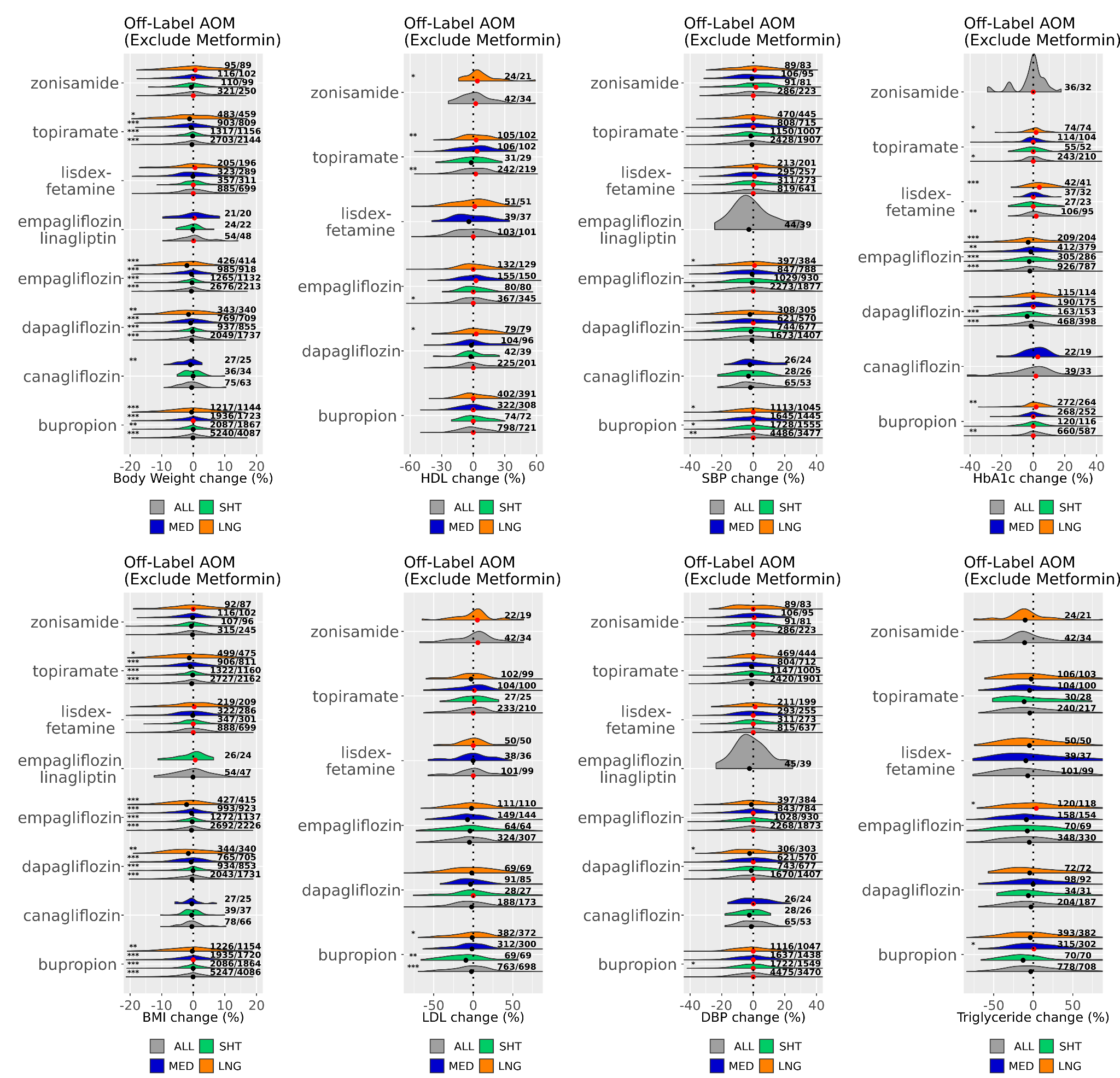

**b**

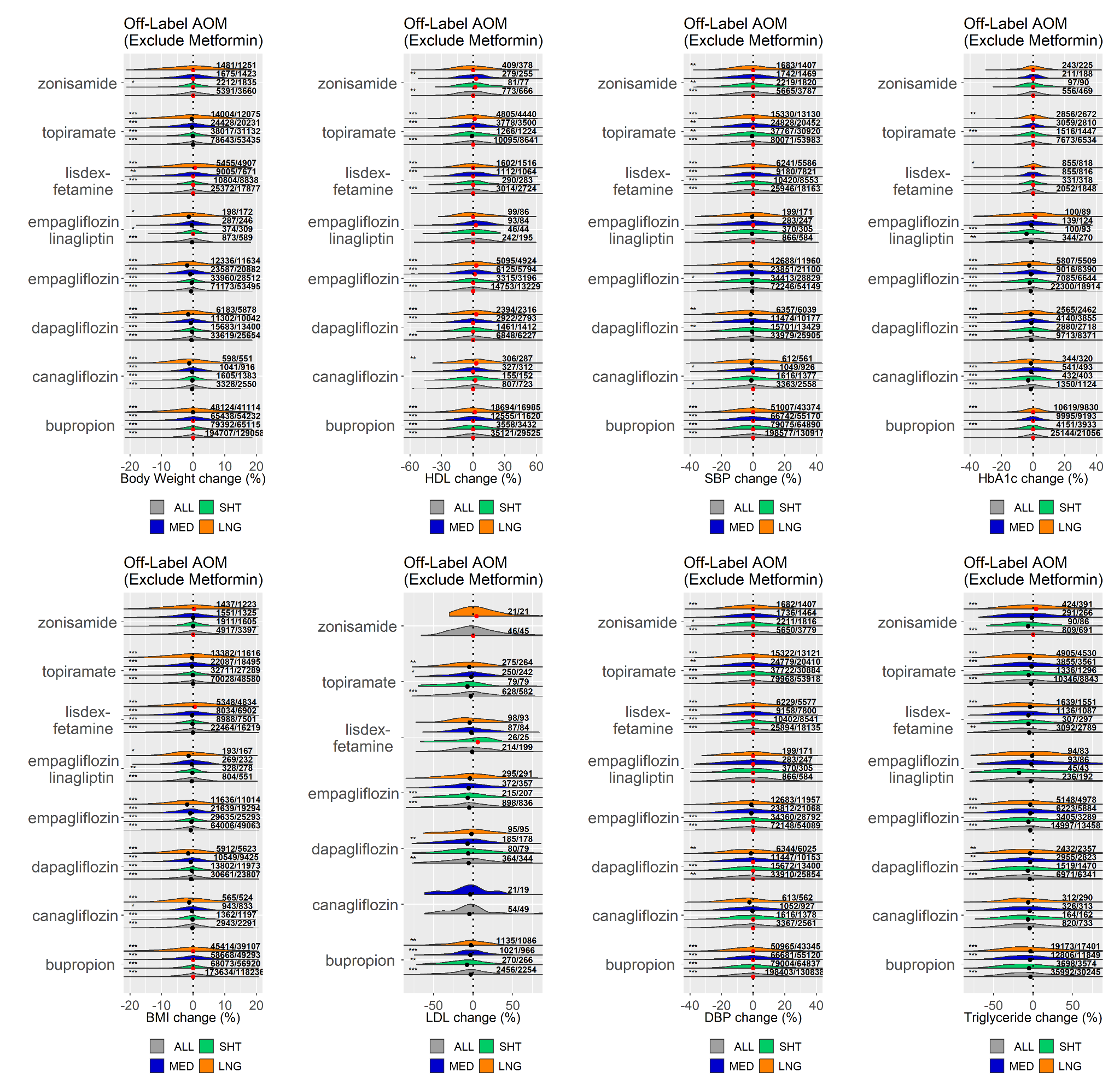

**Supp Figure 4** Distribution of percentage changes in obesity related metabolic metrics after various lengths (SHT: Short-term; MED: Medium-term; LNG: Long-term) of O-AOM (exclude metformin) exposure for a) UT-Physician cohort, and b) COSMOS cohort. Result displayed for each drug and category with a minimum of 20 observations. Black and red dots depict median values <0 and ≥0, respectively. Numbers on the right of each distribution shown as (number of observed records/number of unique patients). The asterisks on the left denote significance (p$\leq$0.05 (*), p$\leq$0.01 (**), and p$\leq$0.001(***)) of two-tailed z-test against the null hypothesis of mean value 0.

**a**

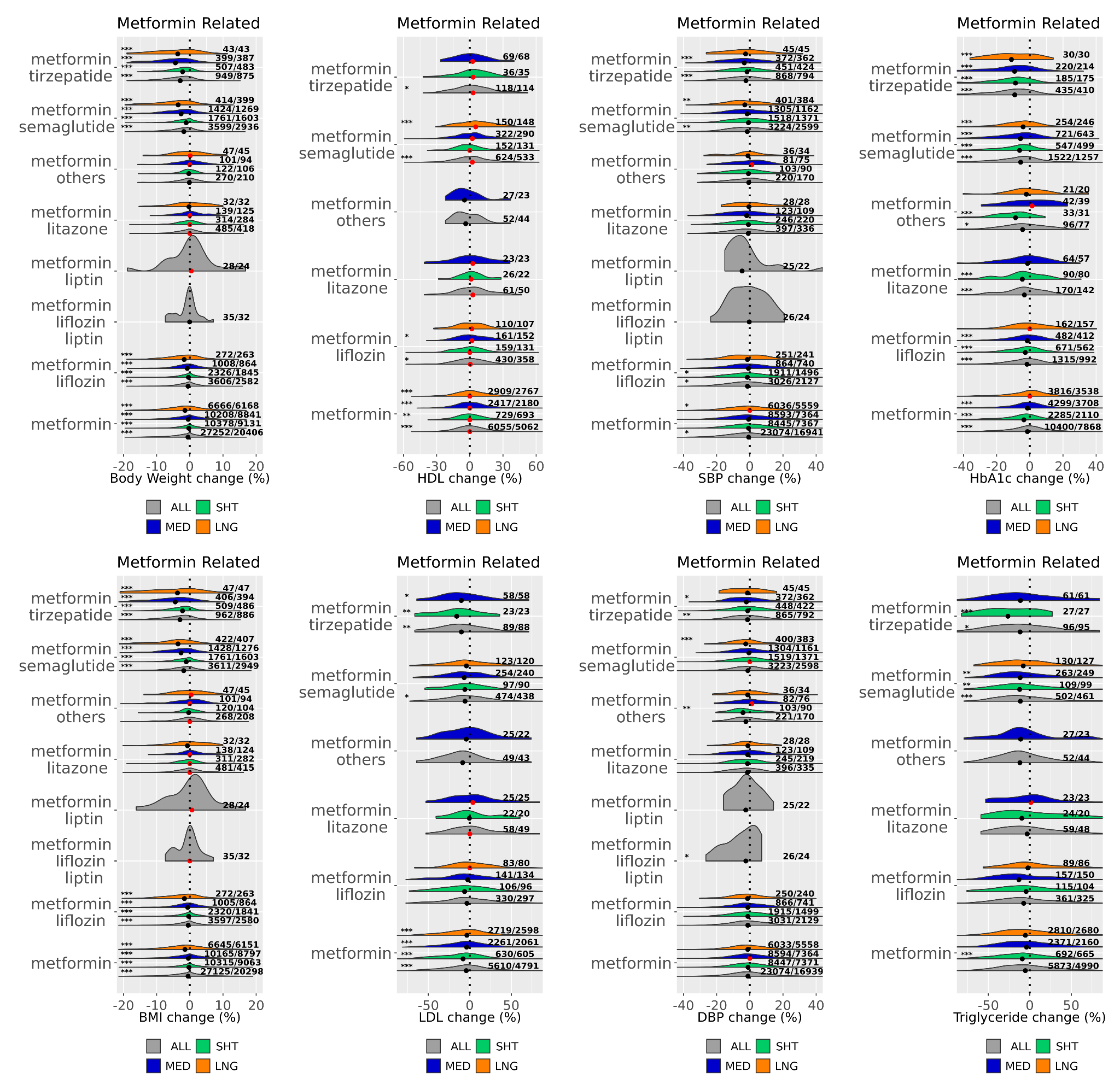

**b**

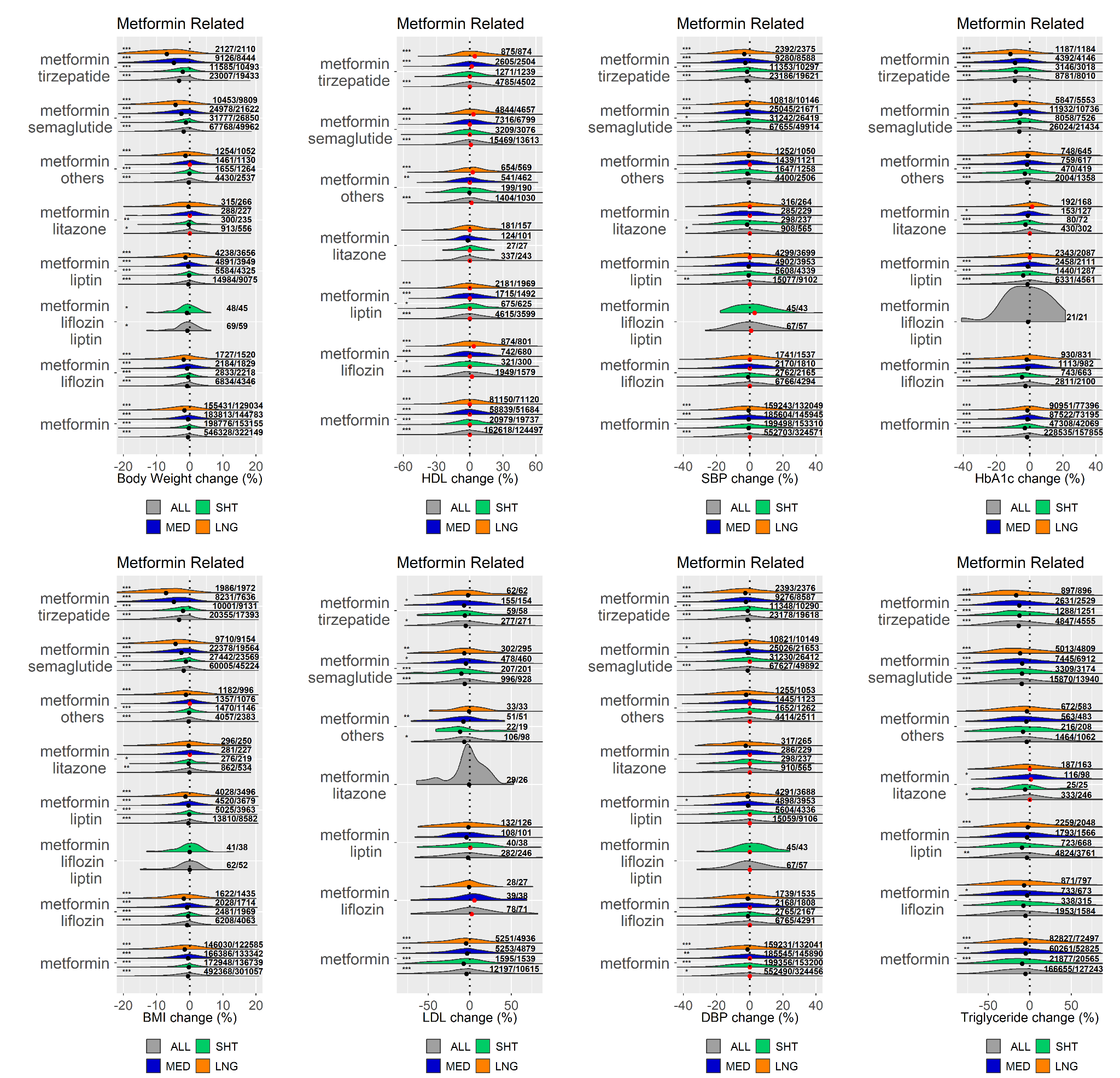

**Supp Figure 5** Distribution of percentage changes in obesity related metabolic metrics after various lengths (SHT: Short-term; MED: Medium-term; LNG: Long-term) of exposure to metformin related AOMs for a) UT-Physician cohort, and b) COSMOS cohort. Result displayed for each drug and category with a minimum of 20 observations. Black and red dots depict median values <0 and ≥0, respectively. Numbers on the right of each distribution shown as (number of observed records/number of unique patients). The asterisks on the left denote significance (p$\leq$0.05 (*), p$\leq$0.01 (**), and p$\leq$0.001(***)) of two-tailed z-test against the null hypothesis of mean value 0.

**a	b**

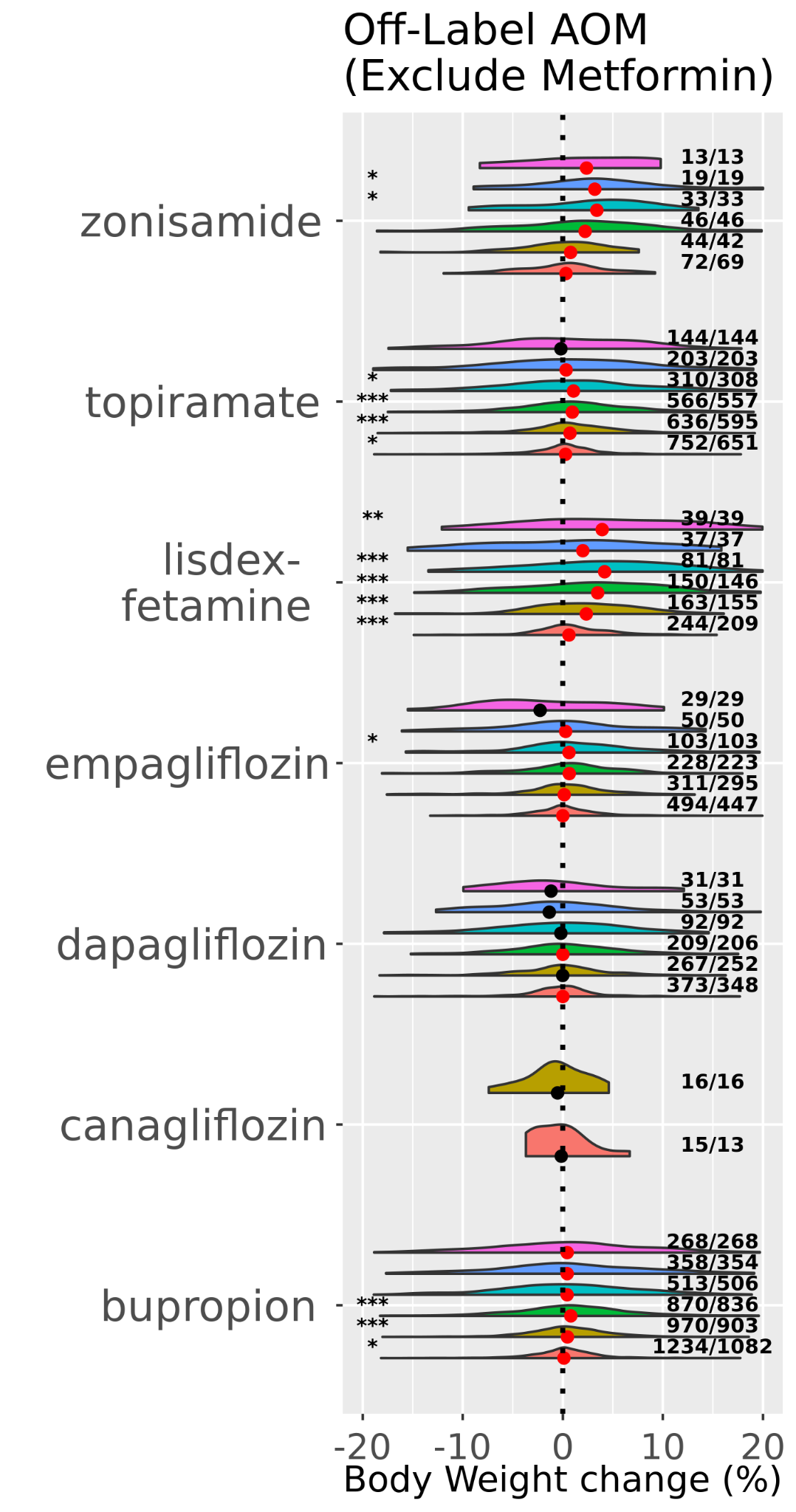

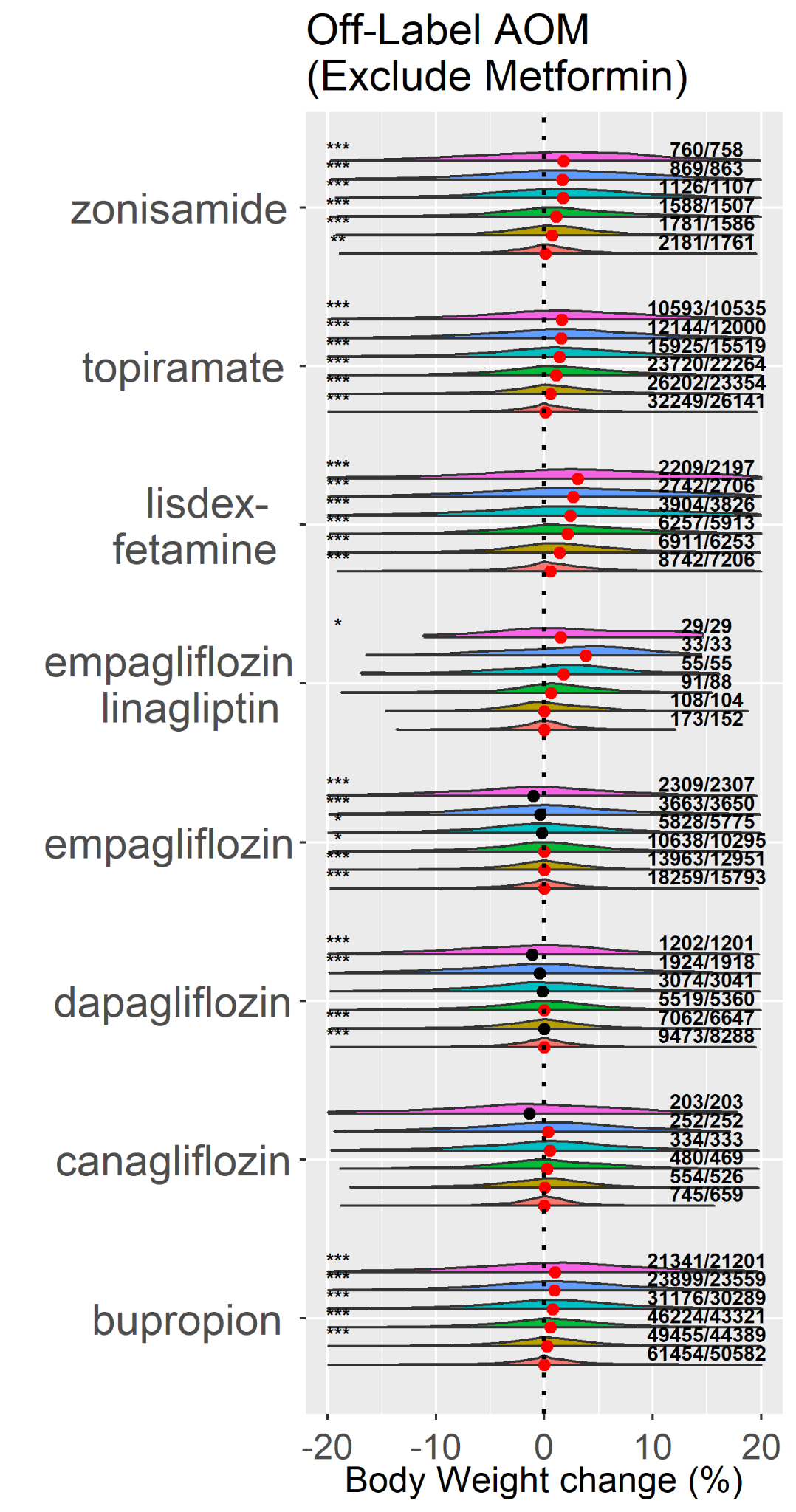

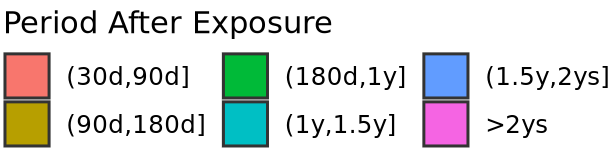

**Supp Figure 6** Body weight changes after discontinuing O-AOMs (exclude metformin), using body weight at the end of exposure as reference weight. a) UT Physician cohort. b) COSMOS cohort.

**a	b**

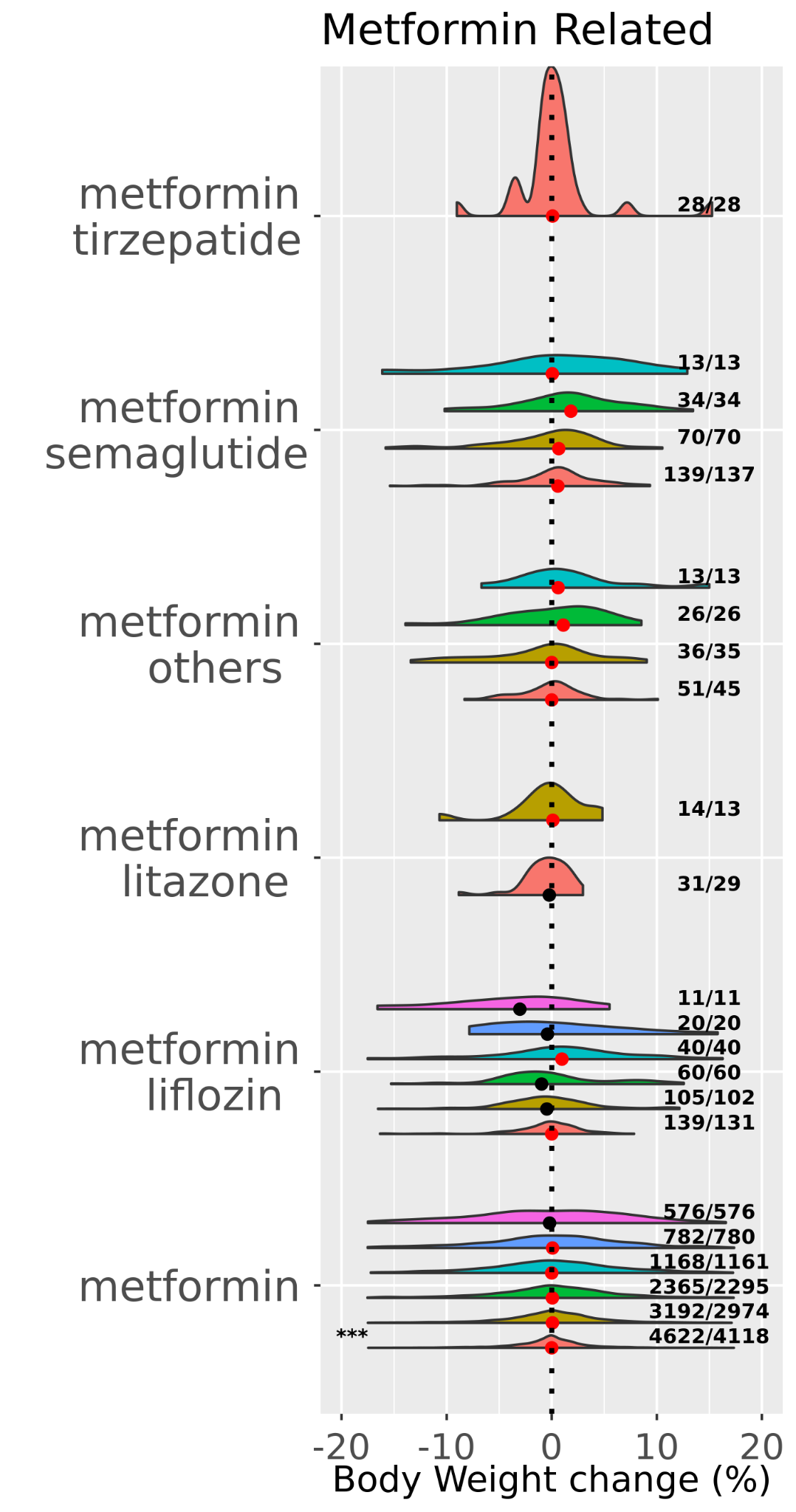

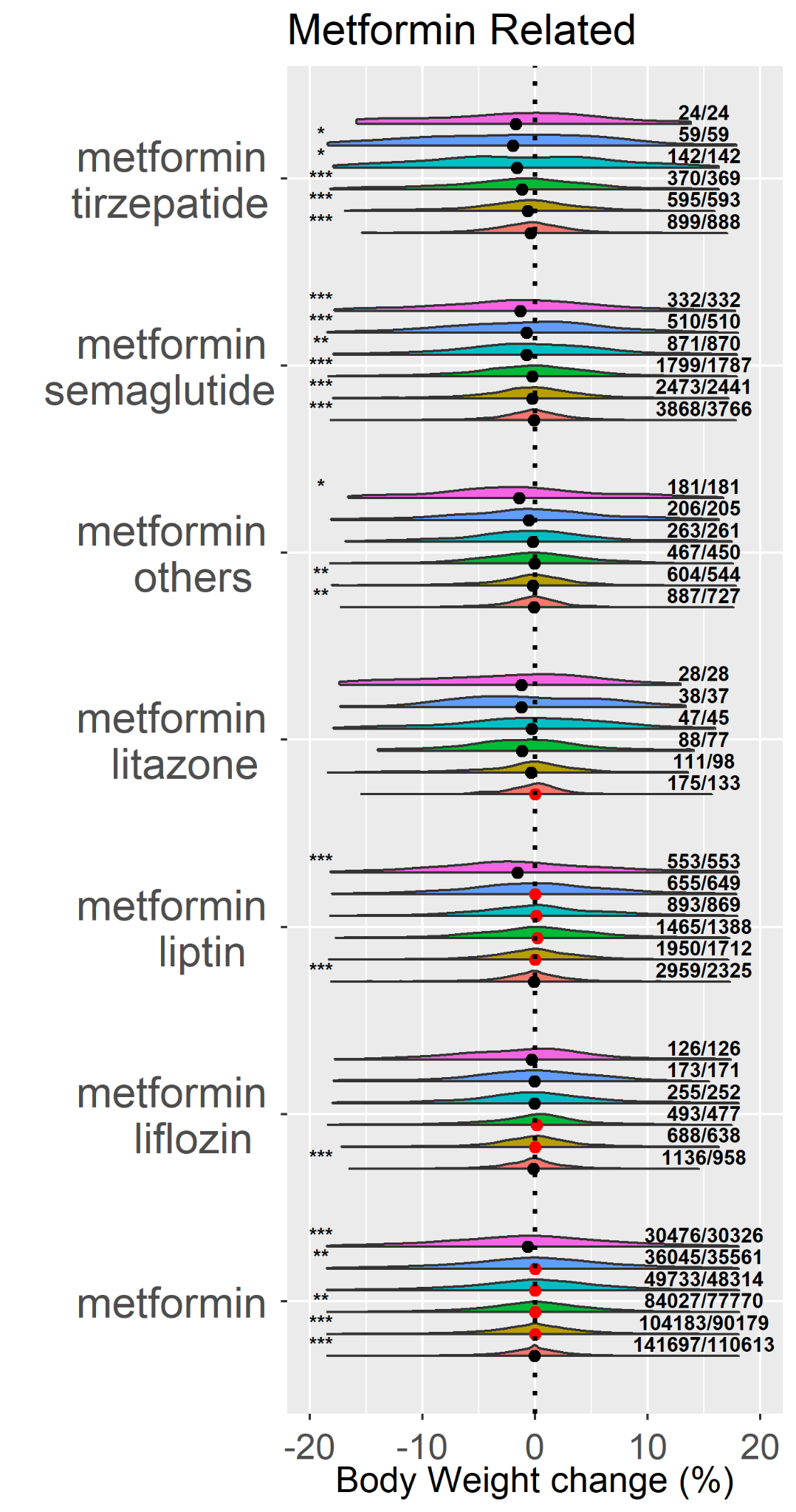

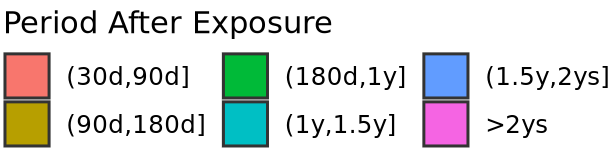

**Supp Figure 7** Body weight changes after discontinuing metformin related AOMs, using body weight at the end of exposure as reference weight. a) UT Physician cohort. b) COSMOS cohort.
